# Identifying barriers and potential solutions to improve equitable access to community eye services in Botswana, India, Kenya, and Nepal: a rapid exploratory sequential mixed methods study protocol

**DOI:** 10.1101/2024.03.07.24303867

**Authors:** Luke Allen, Sarah Karanja, Michael Gichangi, Sailesh Kumar Mishra, Shalinder Sabherwal, Keneilwe Motlhatlhedi, Oathokwa Nkomazana, David Macleod, Min Kim, Jacqueline Ramke, Bakgaki Ratshaa, Malebogo Tlhajoane, Ari Ho-Foster, Nigel M. Bolster, Abhishek Roshan, Mohd Javed, Matthew J. Burton, Andrew Bastawrous

## Abstract

**Introduction:** Evidence suggests that certain groups face substantial barriers to accessing eye care services. This study seeks to explore barriers and potential solutions as perceived by members of the population groups who are least able to access care in the context of four national eye screening programmes. We aim to use rapid yet robust mixed methods that allow us to identify generalisable findings and testable service modifications to improve equitable access to care.

**Methods and analysis:** This is a multi-phased exploratory sequential mixed methods study. First, we will conduct interviews with people purposively selected from the sociodemographic subgroups with the lowest odds of accessing care within each screening programme. Taking a phenomenological approach, we will explore their perceptions of barriers and potential service modifications that could boost attendance at eye clinics among people from these ‘left behind’ groups. We will use a deductive analytic matrix to facilitate the rapid analysis of qualitative data. Space will be made for the inductive identification of themes that are not necessarily captured in the framework. Sample size will be determined by thematic saturation. Next we will conduct a survey with a representative sample of non-attenders from the same left behind groups, asking them to rank each suggested service modification by likely impact. Finally, we will convene a multistakeholder workshop to asses each service modification based on ranking, likely impact, feasibility, cost, and potential risks. The most promising service modifications will be implemented and evaluated in a follow-on randomised controlled trial, the methods for which will be reported elsewhere.

**Ethics and dissemination:** This project has been approved by independent research ethics committees in Botswana, Kenya, India, Nepal and the UK. We will disseminate our findings through local community advisory boards, national eye screening meetings, in peer-reviewed journals, and at conferences.

**Strengths and limitations of this study:**

- We have developed a bespoke rapid qualitative approach that is designed to deliver rich and robust data with speed and relatively low costs. Our approach is based on a prior scoping review of rapid methods.
- By using mixed methods we are able to move from rich data to statistically generalisable findings that can be implemented across four national programmes.
- Our project is embedded withing real-world programmes and will deliver actionable intelligence directly to policymakers, programme funders, and programme implementers.
- Our work places the experience and perspectives of ‘left behind’ groups at the very centre of programmatic quality improvement. This protocol has benefited from the active engagement of lay representatives in each of the four countries.

## Background

Universal Health Coverage (UHC) has been described as the core of the health-related Sustainable Development Goals.^1,2^ As such, boosting access to community-based services has become an important global health priority.^3,4^ Our research team is studying access to eye services in screening programmes that use Peek Vision systems in Botswana, India, Kenya and Nepal (Box 1).^5^ These large screening programmes are identifying hundreds of thousands of children and adults who need glasses, cataract surgery and other cost-effective, life-changing interventions. However, internal data show that only 30-50% of those identified with a need are able to access local treatment outreach clinics, even in programmes where treatment is free. These access figures align with those from other eye services in low-and middle-income countries (LMICs)^6–11^ and with a 2018 review that found mean outpatient clinic attendance to be approximately 50% across a wide range of services and settings.^12^ The ‘central transformative pledge’ of the Sustainable Development Goals is to ‘leave no one behind’, and UN Member States have pledged to identify ‘left behind’ groups and ensure that services ‘reach the furthest behind first’.^13^ We want to ensure that eye care programmes are identifying inequalities in access, engaging with ‘left behind’ groups to understand the specific barriers they face and exploring potential service modifications that would help to improve access.

**Box 1: Peek-powered eye screening programmes**

Peek is a social enterprise that spun out from The London School of Hygiene & Tropical Medicine (LSHTM). Peek have developed a rigorously validated eye screening app that is used in tens of low- and middle-income countries to enable non-specialist teams to perform large-scale community screening programmes.^14–18^ These screening programmes follow two main formats. First, in mobile programmes, a small team works its way through an entire population by sequentially screening children in schools and/or communities in village meeting points or by going house-to-house. An example is Kenya’s Vision Improvement Project that has already screened over a million people. The other type of programme is static, where primary care teams within a given geographic catchment are trained to use the app and then screen patients opportunistically as they present to the primary care facility with other health problems. An example would be the health posts trained to use Peek in Rajbiraj, Nepal. In both cases, screeners use the Peek app to deliver ‘tumbling E’ vision acuity assessments, identifying those whose vision falls below a pre-determined threshold. These positive cases are then referred to local triage and treatment outreach clinics where they are re-assessed by eye professionals and offered eye medication, spectacles, or onward referral for specialist care as required. Peek also provides the patient referral and flow management software that tracks patients through these systems, and can identify 100% of patients who do not attend. Peek is collaborating with LSHTM, the Botswana Ministry of Health, the Kenyan Ministry of Health, College of Ophthalmology of Eastern Central and Southern Africa, Nepal Netra Jyoti Sangh, Shroff Eye Centre, Dr Shroff’s Charity Eye Hospital and the University of Botswana to improve attendance rates and improve equity in screening programme outcomes.

### Literature review: methods to assess barriers to access and potential solutions

#### Whose perspective do we want to hear?

Across all health service research, efforts to understand and redress barriers to access have disproportionately focused on eliciting the opinions and perspectives of ‘experts’ and service providers at the expense of affected people and communities.^19^ Grounding elicitation work in the experiences and perceptions of service users and non-attenders is important both for ethical reasons^19,20^ and because their perceptions often differ to those of service providers.^21,22^ Whilst elicitation studies from the field of eye care have largely been alive to this fact, there are still major issues: the approaches used to explore peoples’ perceptions have been disproportionately based on the use of closed-questions and surveys, or use under-theorised and poorly described qualitative methods.^7,9,21–25^

#### Quantitative vs qualitative approaches for exploring barriers and solutions

The literature on barriers to accessing eye care is dominated by findings from in-person surveys that have been bolted on to population-based screening studies. These commonly take the form of a single survey item where participants are asked to choose or rank reasons for non-attendance from a pre-selected list of options.^8,9,23,24,24,26–30^ This is also the approach used in *Rapid Assessment of Avoidable Blindness* (RAAB) surveys – of which over 300 have been conducted in more than 80 countries.^31^ In our review of the literature, we only found two studies that provided a rationale for the list of barriers that they present to participants: Marmamula et al. asked participants in South India to rank 15 barriers that had been generated by previous focus group work.^32^ However, none of the focus group participants were intended service beneficiaries or people with lived experience of trying to access eye care (all were service providers, public health experts, and researchers).^33^ Furthermore, whilst the people responding to the final survey all had some form of vision impairment, they had not necessarily ever been referred to a service, which may explain why ‘lack of felt need’ and ‘lack of awareness’ were the most frequently selected barriers. Sengo et al performed a literature review and interviewed 25 people in Mozambique with vision impairment to identify which barriers should be used in a wider survey.^34^ However, the exercise was inadequately described and the authors do not provide any detail on how the qualitative data were analysed.

Almost all surveys use a familiar list of barriers that commonly recur in qualitative studies, including costs, distance, lack of trust, communication challenges, fear, scheduling issues, lack of awareness, lack of a chaperone, and low priority accorded to the issue (Box 2).^7,9,21–23,25–28,32,34,35,35,36^ The main limitation in using surveys with these preselected items is that other important factors may be at play in a given population, but it is impossible to ascertain what they are without using open questions.^37^ Methods to elicit these barriers do not have to be particularly sophisticated: even though Sengo et al. appear to have used fairly crude qualitative methods, their study still uncovered important issues including overcrowding in the local hospital, self-medication, and the use of spectacles bought on the street.^34^ Similarly, while the method outlined by Marmamula et al. to interview 199 elderly non-attenders provided no reference to theory, no underlying framework, and no detail on the analytical approach, the work proved vital, with two thirds of respondents citing novel barriers including lack of family consent and the adverse impact of other health conditions.^38^ These factors would not have been elicited from participants through a standard survey.

**Box 2: Commonly cited barriers to accessing eye care**

- High costs
- Distance or transport issues
- Low trust in service providers
- Low perceived service quality
- Poor service communication
- Fear
- Scheduling conflicts or other obligations
- Low awareness of available services
- Lack of a chaperone
- The perception that vision impairment is not a significant impediment to function

When are *any data* better than *no data*? Poorly designed qualitative studies can lead researchers to the wrong- and sometimes harmful conclusions, just as ‘flying blind’ without any understanding of the issues faced by service users can lead managers to introduce well-meaning ‘improvements’ that carry negative unintended consequences. We would argue that using appropriate, theory-driven qualitative methods with a sensible sample and well-described methods is actually a very low threshold to clear and can add real value at low cost in settings where the alternative is not using any open questions at all.

#### Previous qualitative studies that have examined access to eye care

In reviewing the eye care literature, we have found six examples of relatively well-conducted and well-reported studies that have methods designed to explore perceptions of barriers and potential solutions. Ahmad et al. used an open-ended survey question and content analysis to identify barriers to accessing eye care among the general population in Karachi, Pakistan. Unsurprisingly, given the population included, low perceived need was a major reason for not seeking care, however issues around health beliefs and cultural attitudes were surfaced that represent important issues for local health teams to engage with.^39^ Zabeck et al. used structured telephone interviews to explore barriers to access among 28 Americans who had become blind. Using a constant comparative approach they found that social support structures and personal readiness to change were important factors for some people, alongside familiar themes of geographic access and low trust in providers.^40^ Elam and Lee conducted content analysis on data from four focus groups with American community members at risk of not attending eye services. Issues around health insurance, racism, unfriendly service at the clinic, and procrastination supplemented familiar themes of cost, trust, and fear.^35^ Kulkarni and colleagues conducted in-person interviews with transgender people and sex workers with vision impairment in Pune, India, followed by focus group discussions with service providers. Their interview topic guide used deductive (i.e. pre-identified) themes to structure the questions, but also made space “to identify previously unexplored domains”. It appears that the provider focus groups were conducted in parallel in order to triangulate findings from the interviews. This approach was also used in studies led by Owsley and Okoye; both triangulated interview data from the target population with the perspectives of service providers, and Okoye et al also engaged with policymakers.^21,22^

#### Which population should be sampled?

Whilst most eye care studies that assess access have sampled participants from either the general population or the population of intended service beneficiaries, three studies have specifically engaged ‘non-attenders’ (we note that this term is not perfect as it implicitly places responsibility for access onto users rather than services). It is likely that those who have been diagnosed with an eye condition; referred; and not managed to access those services will have greater insight on the barriers to access and potential solutions than members of the general population who do not have this lived experience. Chou et al used a survey with pre-selected items to elicit reasons for non-attendance,^25^ but Gower and colleagues used semi-structured telephone interviews which enabled participants to cite barriers that the researchers might not have considered *a priori*.^7^ Similarly, Marmamula used in-person semi-structured interviews to elicit reasons for low eye clinic access among elderly care home residents.^38^

#### Theory

Very few of the qualitative studies that we found grounded their analyses in theory or a conceptual framework. Whilst there are many different conceptual frameworks on generic barriers to accessing services,^41–43^ we are not aware of any that have been developed for eye care beyond the Australia-focused tripartite division of ‘predisposing’, ‘enabling’, and ‘need’ characteristics described by Keefe et al in 2002.^44^ Despite the breadth of eye service utilisation studies that have been conducted in the past two decades, it seems that it is rare for quantitative or qualitative eye care studies to use theory to inform the design of data collection activities or guide interpretation of findings. Positively, unlike healthcare access research from other fields, approaches that are grounded in eliciting the views of people and communities (as opposed to ‘experts’) are the norm, but these disproportionately sample form the general target population, rather than those with lived experience of being unable to access care.

## Aim

In this study we aim to develop a rapid, theory-based, scientifically robust approach that can be used to elicit barriers to accessing eye care services and potential solutions through engagement with ‘non-attenders’ from sociodemographic groups that experience the lowest overall access rates when referred from screening programmes. We intend to use this approach in eye screening programmes in Botswana, India, Kenya, and Nepal and then apply the findings within the same services with the ultimate aim of improving equitable access to care. Findings from one programme will not be applied to the others, although learning will be shared across sites. All four national screening programmes run on software provided by Peek Vision.

## Objectives

1. In each country, conduct interviews with people from left behind groups who have not been able to access clinics to explore barriers and potential solutions.
2. In each country, conduct phone interviews with a representative sample of people from left behind groups, asking them to rank each of the mooted solutions.
3. In each country, convene the programme funder, programme implementing team, community representatives, and national eye care policymakers at a workshop to review the ranked solutions and select one or more for implementation and evaluation.

### Programme-specific requirements

The nature of the screening programmes imposes a methodologically challenging set of requirements. Given that some Peek-powered programmes screen entire regions in a matter of months, the approach that we use must be able to deliver service modifications rapidly enough to benefit a reasonable proportion of the remaining intended beneficiaries; ideally within weeks-to-months. Next, rather than presenting participants with a pre-selected list of barriers and service modifications and then asking them which are most important, we want to use open questions that allow participants to use their own words to identify issues and approaches that the research team may not have necessarily considered. We recognise that coding and interpreting these responses requires time – however speed is a key objective to ensure feasibility when running at large scale on tight resources. Peek is keen for its programme partners to use any resultant methods that can improve referral uptake, but the cost of these research activities will ultimately be borne by programme funders and will likely be offset by a reduction in the total number of people screened. As such, there is considerable pressure to keep the overall costs as low as possible. A related constraint is that the elicitation approach will only have access to a small number of staff with basic research training. We note that the availability of experienced qualitative and mixed-methods health system researcher staff is low in almost all of the LMICs where Peek-powered programmes operate.^45,46^ Next, as stated above, we want to base decisions on the experiences and perspective of those directly affected; people who have been identified with an eye need and referred, but who have not been able to access services. Furthermore, we aim to focus on the needs of the sociodemographic group with the worst access to care (‘reaching the furthest behind first’ ) so that any improvements disproportionately benefit these groups, thereby improving equity (in line with the idea of proportional universalism). Finally, despite being rapid, inexpensive, non-prescriptive, equitable, and primarily conducted by non-experts, we are committed to using robust methods to deliver valid, non-tokenistic findings. This is vital in order to inform programmatic changes that stand a chance of improving access rates (Box 3).

**Box 3: Our improbable wish-list**

We want to develop a rapid elicitation tool that:

- Can deliver a set of barriers and potential solutions within weeks-months
- Uses open questions rather than a pre-defined list of response options
- Provides barriers and potential solutions that are generalisable
- Gathers data from non-attenders from sociodemographic groups with the lowest attendance rates within each programme
- Can be largely conducted by non-experts, albeit with expert supervision
- Is inexpensive
- And is methodologically robust

## Approach

### Philosophical paradigm

Our aim requires methods that span the space between constructivist and positivist philosophical paradigms.^47^ Whilst the task of seeking to understand perceptions of barriers and solutions is primarily phenomenological, we intend to generalise the findings (i.e. make statistical inferences) and develop service modifications that will be applied across entire programmes within each country. To traverse this philosophical rift we will use a pragmatist paradigm, originally advanced by Charles Sanders Peirce.^48,49^ Pragmatism holds that ‘truth’ is determined by practical application and consequences, and it is agnostic on the type of research techniques used as long as they answer the research question.^48,50^

### Undergirding theory

There are a large number of conceptual frameworks on access to health services.^41,43,51–55^ As our ultimate aim is to elicit ideas for ways of improving services to boost equitable access, we have elected to use the popular model developed by Levesque and colleagues (Figure 1)^43^ that divides factors into those pertaining to services and those relating to potential service users. We want to focus our analysis on areas that we are most able to change i.e. the structure, staffing, organisation, and communications of eye services, in contrast to user characteristics like social support networks, assets, and health literacy which are important but much harder for us to influence.

The Levesque framework is based on the findings of a systematic review that identified five determinants; approachability; acceptability; availability & accommodation; affordability; and appropriateness, along with corresponding abilities to perceive, seek, reach, pay for, and engage with services. These factors feed into a process of seeking care that resonates with the Tanahashi framework^56^ and the concept of effective coverage^57^ i.e. access is predicated on a series of steps that include perceiving an initial need, desiring care, seeking out potential providers, traveling to the location at a time that it is open and staffed, and having sufficient resources to be seen. Access only occurs when the requisite supply and demand side elements are in place.

**Figure 1:**
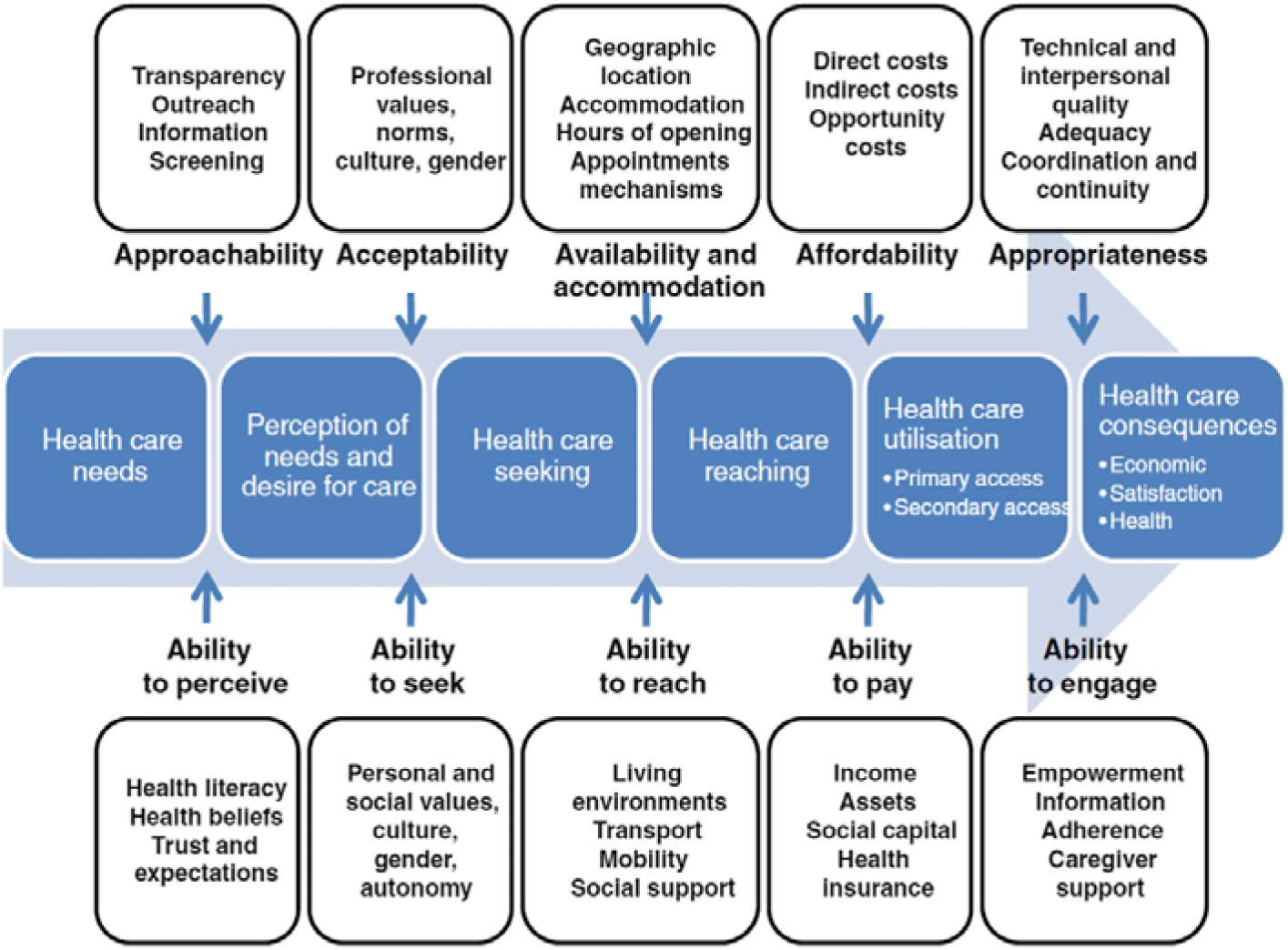
The Levesque framework.

Obrist and colleagues have developed an aligned model with a specific emphasis on ‘analysis for action’ and application in low-income settings.^41^ Their five dimensions; availability, geographic/logistical accessibility, affordability, adequacy, and acceptability (Table 1) overlap with those presented by Levesque, and are supplemented by five types of livelihood assets that determine ability to recognise need and seek out health services: human capital (local knowledge, education, skills); social capital (social networks and affiliations); natural capital (land, water, and livestock); physical capital (infrastructure, equipment, means of transport); and financial capital (cash and credit). The authors note that many of these assets are influenced by macroeconomic and political conditions, climate change, and many other forces over which people have very little control, and are also difficult for service managers to influence directly.^41^

**Table 1:** Obrist’s five dimension of access.

| Dimension | Questions |
| --- | --- |
| Availability:<br>The existing health services and goods meet clients' needs. | What types of services exist? Which organizations offer these services? Is there enough skilled personnel? Do the offered products and services correspond with the needs of poor people? Do the supplies suffice to cover the demand? |
| Accessibility:<br>The location of supply is in line with the location of clients. | What is the geographical distance between the services and the homes of the intended users? By what means of transport can they be reached? How much time does it take? |
| Affordability:<br>The prices of services fit the clients' income and ability to pay. | What are the direct costs of the services and the products delivered through the services? What are the indirect costs in terms of transportation, lost time and income, bribes, and other "unofficial" charges? |
| Adequacy:<br>The organization of health care meets the clients' expectations. | How are the services organized? Does the organizational set up meet the patients' expectations? Do the opening hours match with schedules of the clients, for instance the daily work schedule of small-scale farmers? Are the facilities clean and well kept? |
| Acceptability:<br>The characteristics of providers match with those of the clients. | Does the information, explanation, and treatment provided take local illness concepts and social values into account? Do the patients feel welcome and cared for? Do the patients trust in the competence and personality of the health care providers? |

### Methodology

We require mixed methods that draw on the strengths of both qualitative and quantitative approaches to answer a multi-layered question: what are the main barriers to accessing eye services in each location and what can be done about them?

Qualitive methods deliver rich, descriptive data based on interviews, discussions, and/or observations with a select number of participants who are often purposively chosen because of their specific characteristics. As such, the findings can be transferred to similar cases and contexts, but they are not intended to be generalisable. In contrast, quantitative methods deliver numerical data and - with representative sampling - are able to provide evidence for causality, generalisability and magnitude of effect.^58,59^

We will use a mixed methods approach; starting with qualitative methods to explore non-attenders’ perceptions of the barriers and potential solutions in each setting. We will use the identified themes to develop a unique, user-derived list of potential service modifications within each screening programme. We will then use quantitative methods – a survey - to establish which of these are perceived to be the most impactful through engagement with a representative sample of non-attenders, effectively validating or ‘sense-checking’ the qualitative findings with a larger, representative group. The ranked suggestions for service improvements will then be taken to a multistakeholder workshop where the top-ranked solutions will be considered for implementation based on their likely impact, feasibility, cost, and potential risks.

### Context

This project constitutes the ‘Engage’ element of the broader ‘IM-SEEN’ continuous improvement approach.^60^ It is preceded by activity to gather sociodemographic data from those being screened in each setting and the identification of which groups experience the lowest access rates (Figure 2). The purpose of the current ‘Engage’ project is to gather and prioritise a list of barriers and potential solutions, grounded in the perceptions of left behind groups. A follow-on project will use an RCT to test whether the most promising solution(s) actually equitably improve access to services.

**Figure 2:**
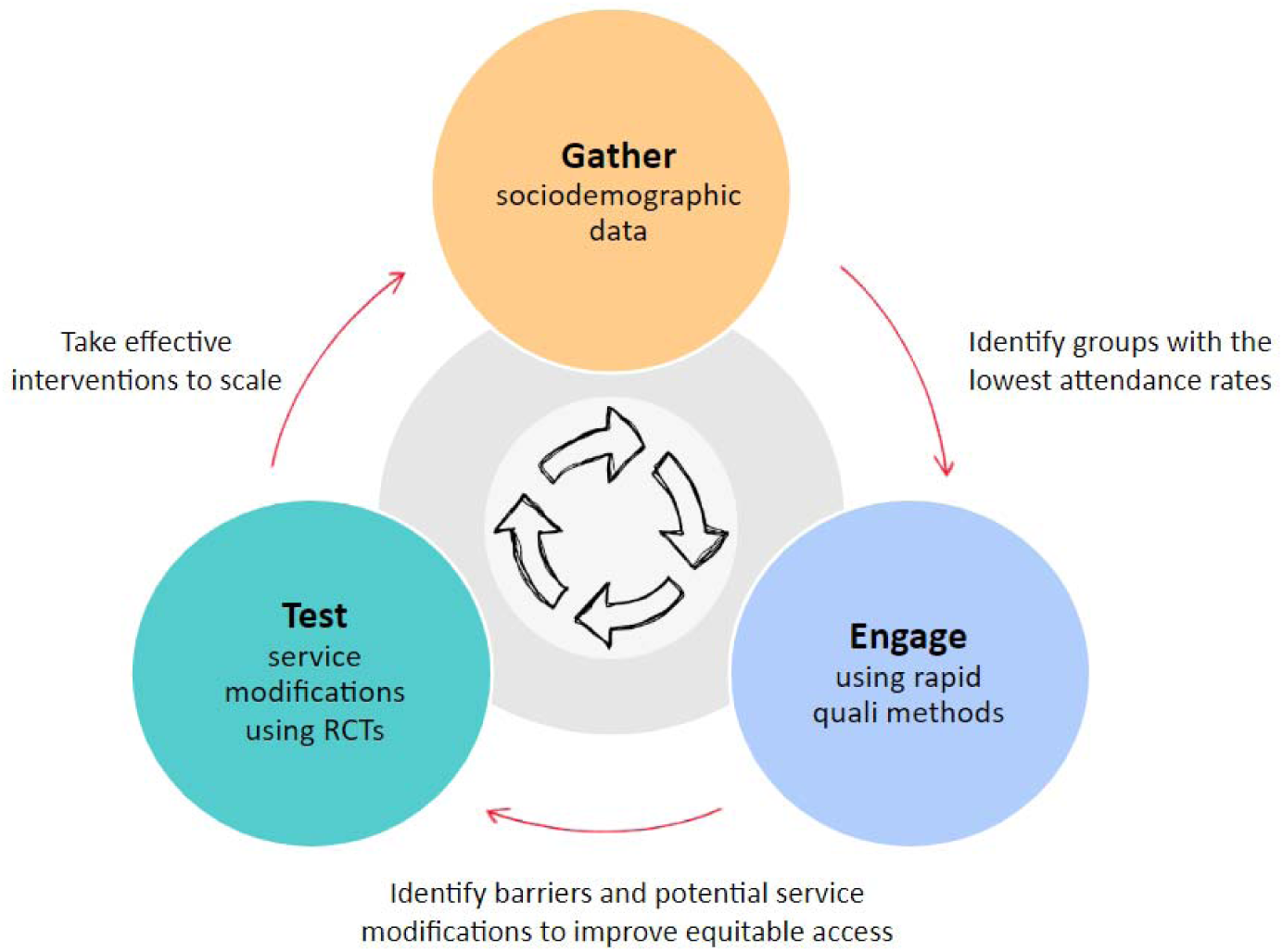
This current project represents the ‘Engage’ component in the wider ‘IM-SEEN’ continuous improvement project.

## Methods

### Summary

We will use a four-stage rapid exploratory sequential mixed methods study design (Figure 3). First, we will conduct telephone interviews with non-attenders purposively selected from the sociodemographic subgroup that has the lowest overall access rate within each screening programme. We will explore their perceptions of barriers potential solutions and compile a long list of all suggested solutions/service modifications. We will discuss the long list with the programme funder and implementer to rule out any suggestions that are felt to be completely unfeasible e.g. providing helicopter transport for everyone who is referred. Next, we will conduct a telephone survey, asking a representative sample of non-attenders from the same left behind group to rank the remaining suggestions by likely impact. Finally, this list of prioritised service modifications will be put to a group of programme funders, programme implementers, community representatives, and eye care policymakers. Participants will review the top-ranked service modifications and select one or more to test based on likely impact, feasibility, cost, and potential risks. The intervention that is perceived to offer the best value according to these criteria will be implemented and evaluated within the context of an embedded pragmatic randomised controlled trial, the methods for which will be reported elsewhere. This approach will be conducted independently in each country. Figure 3 provides an overview of the study elements.

**Figure 3:**
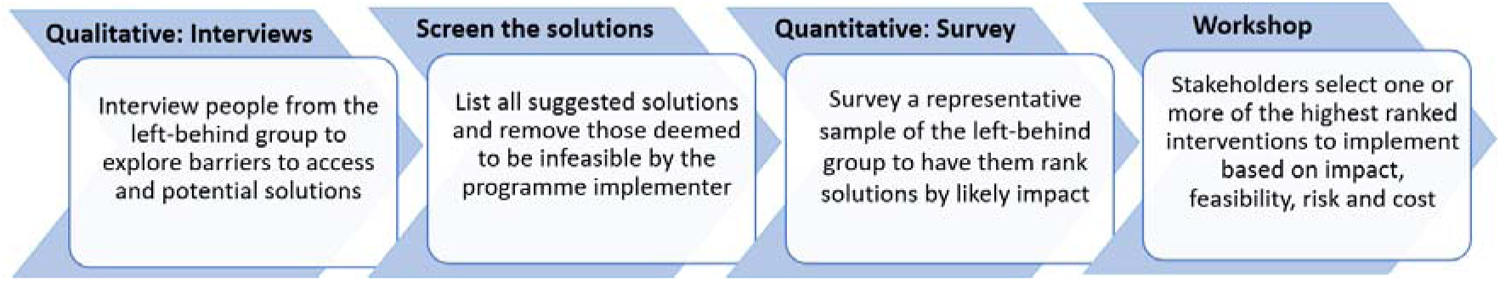
Overview of the sequential mixed-methods approach.

### Developing a rapid qualitative approach

Our study is not the first that seeks to use rapid and low-cost qualitative methods that can be led by less-experienced researchers (early-career researchers and those with basic-rather than postgraduate training) to answer an open question. Rapid methods have been in use for over 30 years, as described by Beebe,^61–63^ Handwerker,^64^ Pearson,^65^ Bentley,^66^ Scrimshaw et al.,^67^ and Johnson and Vindrola-Padros.^68,69^ There are also examples of rapid qualitative studies that have intentionally used teams of less experienced researchers.^70^

Rapid qualitative methods are often used to reduce time and costs, and to improve efficiency, accuracy, and ‘obtain a closer approximation to the narrated realities of research participants‘.^69^ These studies generally take between a few days to a few months, depending on the design, with most taking a couple of weeks to complete.^68,71^ A large number of dedicated approaches have been developed, including ‘Rapid Ethnographic Assessment’,^72^ ‘Participatory Rural Appraisal’,^73^ ‘Rapid Rural Appraisal’,^74^ ‘Rapid Appraisal’ (a form of ‘Rapid Qualitative Enquiry’),^61^ ‘Rapid Assessment Procedures’,^61,67^ and ‘Rapid Assessment Response and Evaluation’.^75,76^

In their review of rapid qualitative methods, McNall and Foster-Fishman identify the following key features: these studies commonly use mixed and multi methods to triangulate data; they tend to be participatory – with representatives of the target population involved in planning and implementation; they are team-based with all members working collaboratively on all aspects of the research process; and they are iterative - with data being analysed as they are collected and early findings being used to guide additional data collection until theoretical saturation is reached.^77^ The authors also note that the central trade-off is between speed and trustworthiness. Vindrola-Padros and Vindrola-Padros identified seven key challenges that apply to all rapid qualitative approaches, summarised in Table 2.^71^

**Table 2:** Risks of rapid research, as described by Vindrola-Padros and Vindrola-Padros^71^.

| Design feature | Potential risks |
| --- | --- |
| Sample size and representativeness | ‘Dependency on most accessible informants and loss of multiplicity of voices.’ |
| Community participation | ‘Local research assistants are not always available, have the required skills or willingness to take part. Training takes time. Research undertaken by researchers without an anthropological background might limit the quality of the study’ |
| Team-based approach to | ‘Recruitment might be an issue and clear roles in the field need to |
| design, data collection and analysis | be outlined' |
| Brief engagement time | 'Inability to capture changes over time, understand all relevant social and cultural factors at stake, or conflict and contradictions... New researchers might get more attention, but lack familiarity with the study area. Prolonged engagement often increases credibility and internal validity. Prolonged engagement might also lead to stronger relationships between research participants and the field researchers. The rapid study timeframes might not allow researchers to critically analyse the position they play in the field site and their role in the collection and analysis of data.' |
| Governance | 'Time pressures should not deter researchers from undergoing the required governance and informed consent processes.' |

Many of these risks can be met head-on e.g., by obtaining ethical approval and informed consent, thinking carefully about team roles, and purposively sampling from the most marginalised groups. The extent to which community members can or should be engaged is dependent on the study aims and local contextual factors. The greatest challenges are around developing robust findings based on a brief engagement period. Triangulation can help (i.e. using multiple methods or data sources to develop a comprehensive understanding of phenomena (p247^78^) but this limitation renders rapid methods unsuitable for qualitative research projects that require a deep, emic understanding of complex phenomena and issues.

Building on established rapid qualitative analysis, our team has conducted a scoping review to identify rapid approaches that have been specifically used to assess barriers and solutions to improve access to community health services.^79^ We identified a number of innovative methodological techniques that can be used to minimise the length of time between data collection and implementation of the final set of findings. Many of these design features are best suited for deductive framework analyses where participants’ experiences are sought in relation to a clearly defined a priori research question, in our case; ‘what stopped you attending and what could be done about it?’

In line with findings from a broader review of rapid methods,^69^ we found that many approaches focused on eliminating or expediting the transcription phase, either by performing simultaneous data collection and analysis, or by coding data directly from audio. This is a common design feature of studies that use ‘RAP’ sheets (*Rapid Assessment Procedure* data templates): data collectors enter quotes and/or open codes into analytic matrices during the interview or afterward, working directly from the audio recording.^70^ Clearly this limits the depth and richness of the analysis and makes the approach inappropriate for complex and nuanced qualitative research questions, however many applied research teams have used contemporaneous analysis to elicit meaningful and non-tokenistic findings in contexts where there is a narrow and clearly articulated aim. The few methods studies that have compared these direct coding approaches against coding based on transcripts of the same interviews/focus group discussions found that both approaches generated similar themes with acceptable reliability.^80,81^

In our scoping review, we found that the most commonly used application of direct coding was in entering data into a deductive template during the interview and/or directly afterwards, working from handwritten notes and/or the audio recording rather than a transcript. The loss to analytical power from obviating a written record can be partly offset by having data collectors co-located, which has been shown to lead to informal discussion and analysis through natural debriefing conversations.^70^ Some researchers have formalised this process, holding group meetings directly after data collection to collaboratively summarise, analyse, and interpret findings, such as in the work led by Jalloh.^82^

Many rapid studies seeking to understand barriers to healthcare access make use of deductive templates or matrices to chart data or use ‘one sheet of paper’ techniques to aid rapid analysis and presentation of findings.^82–86^ Miles and Huberman have argued that data reduction, display, and the drawing of conclusions happens simultaneously in qualitative analysis (p10),^87^ and that the use of matrices can drive credibility and trustworthiness.^88^ Whilst the use of a priori codes and/or themes to populate a framework template may save time at the analysis stage and potentially reduce the skill requirement, the burden of work is shifted to an earlier stage of the project rather than eliminated.

A further issue is that deductive approaches are misaligned with the general aim of moving away from pre-selected checklists of potential barriers and making space for affected people to describe the issues in their own words, ideally surprising researchers by describing barriers and potential solutions that had not previously been considered, and by ‘making the familiar strange’.^89^ However, Pope and Mays argue that virtually all qualitative analytic approaches involve a combination of inductive and deductive reasoning, and the use of a deductive framework does not necessarily preclude inductive coding.^47^ They make a strong case for ‘abductive’ reasoning that benefits from the efficiencies of the deductive framework approach whilst “leaving space for more inductive identification of themes and issues not predicted at the outset” (p19).

Based on the lessons learned from reviewing the literature, we aim to adopt several rapid techniques to increase the speed and affordability of our qualitative research element, detailed below.

### Interviews with non-attenders or their proxies

#### Recruitment and sampling

Participants in Peek-powered screening programmes operating in Botswana, Inda, Kenya, and Nepal provide their name, a contact number and - if they consent - data on approximately ten sociodemographic domains including age, sex, education, income, assets, and health status (the unique lists for each national programme and selection processes have been detailed in a previous IM-SEEN publication^90^). Peek has consent procedures and agreements that enable these data to be shared with our embedded research team. In each country we will conduct quantitative equity analyses to identify which sociodemographic characteristics are most strongly associated with non-attendance in each programme. This work has already been completed in Meru, Kenya, where we found that younger people, males, and those working in sales, services, and manual jobs were the least able to access care. In our intersectional analysis we found that only 14% of young men who worked in sales, services, and manual jobs accessed clinics in comparison to 50% across the entire referred population.

In line with the global health principles of equity and health for all, in each setting we will purposively engage with the sociodemographic groups that are found to experience the lowest access rates. We will purposively recruit people who have been referred but not accessed care within two weeks of their appointed date from the left behind subpopulation.

We will have the phone numbers for every person who did not access care from the left behind subpopulation. We will generate a spreadsheet that contains each person’s name, unique study ID number, phone number, and screening date. We will order the names randomly, using a random number generation function in R or Excel, and then work down from the top of the list.

Our sample size will be determined by the point at which we reach thematic saturation. Empirical evidence suggests that the majority of all themes and concepts emerges within the first 5-6 interviews^91,92^ and that saturation is usually reached within 9-17 interviews when conducted among a relatively homogeneous population.^93,94^ We will use Guest and colleagues’ approach to assessing saturation, using a prespecified base size (i.e. a minimum number) of 12 interviews, followed by runs of two interviews and a 0% new information threshold. In other words, we will stop conducting new interviews once no new themes emerge after two interviews in a row, with a minimum sample size of 14 (‘12+2’). We will budget conservatively for 20 interviews in each location.

##### Data collection

Small teams of data collectors will conduct interviews in each country. All data collectors will have at least basic training in qualitative methods but will not necessarily be full-time qualitative researchers. Where possible we will recruit, and train lay members from the target population to assist with data collection. All data collectors will be fluent in the language(s) spoken by the target population.

We will use semi-structured telephone interviews, directly exploring participants’ views of the issues that prevented them from attending clinic and the potential service modifications that they feel would have enabled them to attend. We will call potential participants and explain the study, and then seek recorded audio consent. All interviews will be conducted in the participant’s own language.

Whilst face-to-face interviews probably offer richer data in comparison with telephone interviews, we have opted for the latter on the basis of feasibility. Peek do not collect people’s home addresses, and even if we did have this information, the national screening programmes cover extremely large areas, meaning that it might take weeks of travel to conduct the interviews. In contrast, multiple phone interviews can be conducted each day, with much lower costs, whilst avoiding the personal safety risks to data collectors that come with extensive travel. A number of methods papers have argued that qualitative findings do not vary significantly between telephone and in-person modalities.^95,96^

We will try to contact each interviewee three times, calling at different times of the day. If we are unable to reach them, we will move down the randomly sorted list and try the next non-attender. Interviews will be audio recorded. The recording will include the participants’ unique identifier, the consent process, and – if given – confirmation of consent to participate. The following interview items will be used:

**Barrier elicitation questions**
• In your own words, can you talk me through why we didn’t see you/your child at that clinic?
**Probing questions**
• Are there any other factors that prevented you/him/her from attending?
• Is there anything else you’d like to share?
**Solution elicitation questions**
The last part of the interview is exploring whether there is anything we could do to address these barriers and make it more likely that other people like you/children like [child’s name] will attend in the future.
• So, to start, what would make the biggest difference?
**Probing questions**
• What else would help?
• What other changes could we make to the programme that would make it easier for you/children like [child’s name] or people like you/children like [child’s name] to attend?
• Are there any other specific changes that we could make to the way that the programme or eye clinics run?

### Qualitative Analysis

During the interview, data collectors will note the major barriers and solutions, and the time that they were mentioned. Immediately after the interview has concluded, the data collectors will listen back to the interview recording and navigate to the noted times. They will then type out the full quotes for each barrier or proposed solution verbatim into an analytic matrix, working from the audio recording, with one interviewee per column, and one theme per row.

We have chosen to use this direct data entry approach because it is faster than generating and then working from transcripts, and because the nature of our (relatively simple) research question is more descriptive than explanatory. We have developed a bespoke deductive matrix that is grounded in the access models of Levesque et al.^43^ and Obrist^41^ et al.

### Development of the analytic matrix

We first mapped the Obrist dimensions to the service domains identified by Levesque (Table 3).

**Table 3:** Mapping Obrist’s service dimensions to those described by Levesque et al.

| Levesque dimensions | Levesque descriptors | Aligned Obrist dimensions and descriptors |
| --- | --- | --- |
| **Approachability** | Transparency, outreach, information, screening | N/A |
| **Acceptability** | Professional values, norms, culture, gender | 'Acceptability' – Provider norms and values align with users, ~~trust,~~ patients feel welcomed and cared for |
| **Availability** | Geographic location, accomodation, opening hours, appointment mechanisms | 'Accessibility' –Geographic location, transport options, and time to travel<br>'Adequacy' – the service organiation and opening times meets clients' expectations. Facilities are clean and well kept |
| **Affordability** | Direct-, indirect-, and opportunity costs | 'Affordability' - Direct-, indirect-(including bribes and unofficial charges), and opportunity costs |
| **Appropriateness** | Technical and interpersonal quality, adequacy, **coordination and continuity** | 'Availability' - The service meets clients' needs: enough skilled personnel, products and services correspond with needs and **cover demand**<br>'Acceptability' – ~~Provider norms and values align with users,~~ trust, patients feel welcomed and cared for |

Next, unencumbered by the requirement to begin all descriptors with the letter ‘A’, we selected domain descriptors that we felt captured the essence of each unique element from across the two frameworks (Table 4). We felt that Levesque’s ‘availability’ domain straddled two different concepts; those relating to distance/transport and facilities.

**Table 4:** Drawing out unique service domain terms.

| Service domains | Levesque | Obrist | Unified service descriptor |
| --- | --- | --- | --- |
| Awareness of the service | 'Approachability' - Transparency, outreach, information, screening | N/A | The service provides clear information about what is available to potential beneficiaries in the catchment population |
| Cultural values and health beliefs | 'Acceptability' - Professional values, norms, culture, gender | 'Acceptability' – Provider norms and values align with users | The service norms and values align with those of intended users e.g. around gender interactions or health beliefs |
| Distance and transport | 'Availability' - Geographic location, ~~accommodation,~~ ~~opening~~ hours, ~~appointment mechanisms~~ | 'Accessibility' – Geographic location, transport options, and time to travel | The service is nearby and served by good infrastructure and transport options |
| Facilities | 'Availability' - ~~Geographic location,~~ accomodation, | 'Adequacy' – the service organisation and opening times meets clients' | The facilities are clean, well kept, well organised, and open at predictable and convenient |
|  | opening hours, appointment mechanisms | expectations. Facilities are clean and well kept | times |
| Costs | 'Affordability' - Direct-, indirect-, and opportunity costs | 'Affordability' - Direct-, indirect- (including bribes and unofficial charges), and opportunity costs | The direct costs of care, associated costs, and opportunity costs are all affordable for intended beneficiaries |
| Service quality | 'Appropriateness' - Technical and interpersonal quality, adequacy, coordination and continuity | 'Availability' - The service meets clients' needs: enough skilled personnel, products and services correspond with needs and cover demand<br>'Acceptability' -Clients trust the providers and feel welcomed and cared for | Services are well stocked and staffed by competent staff who are able to meet the needs of intended beneficiaries with warmth and care |

Note: Text in the descriptors that is not relevant for the domain in question has been struck through.

Next we added in the domains that pertain to users, mapping them to the service domains and providing a unified descriptor (Table 5). The Levesque framework identified three areas that do not naturally correspond with service characteristics: themes around the desire to seek care, the capacity to participate in care (e.g. though shared decision making with a clinician or medication concordance), and empowerment and social support.

**Table 5:**
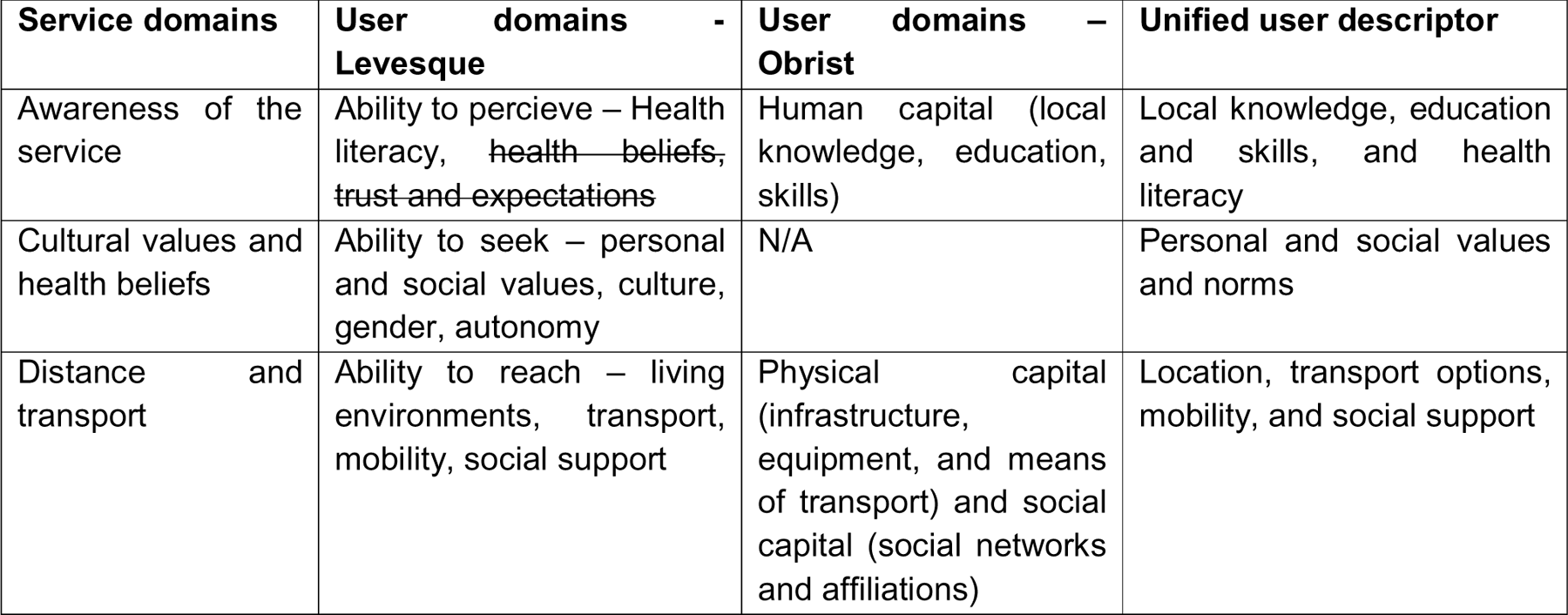
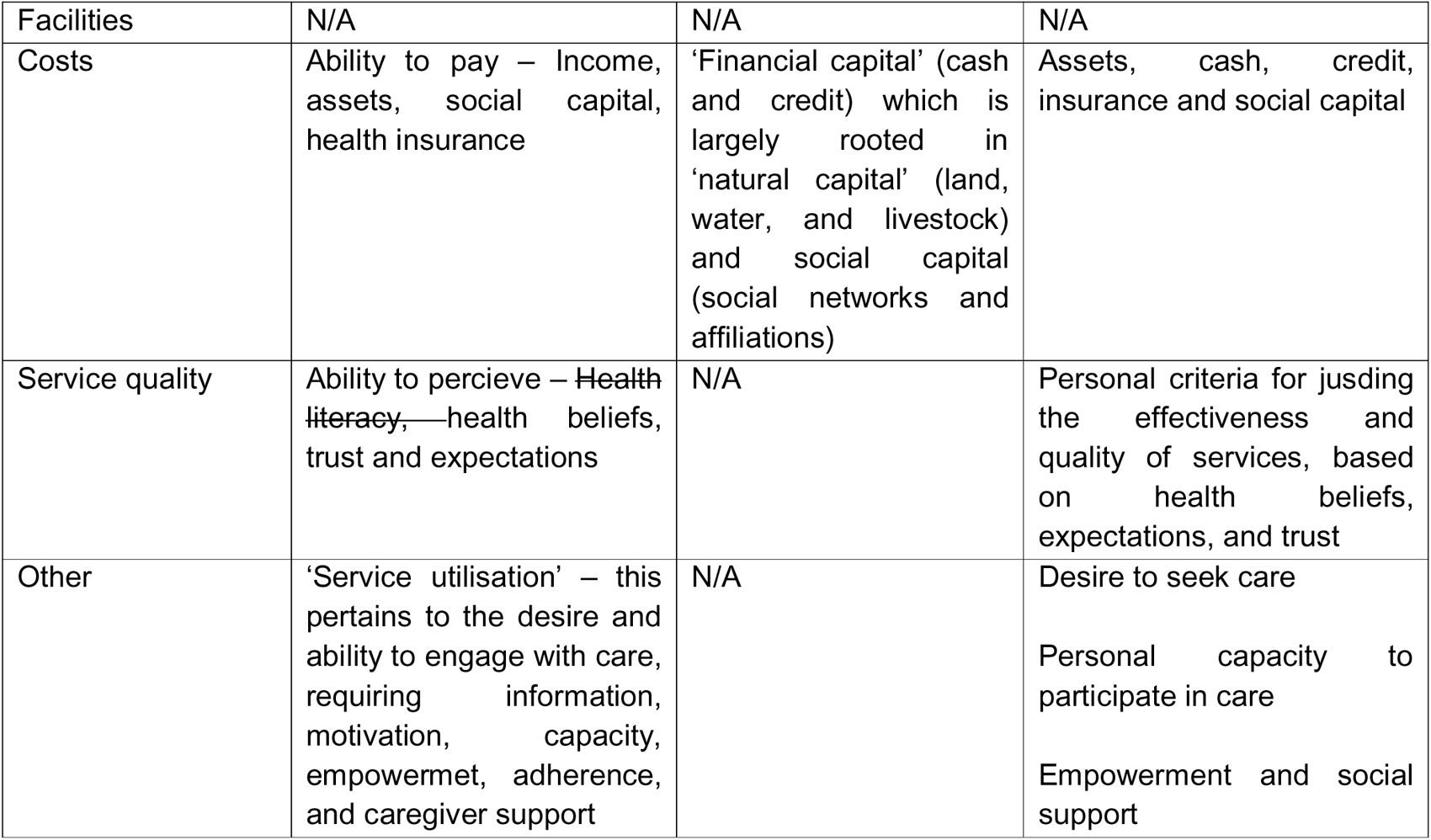
Adding in corresponding service user domains.

Next we mapped the common barriers that were indentified in our literature review of the existing eye care literature (Box 2) to the unified descriptors of service and user domains (Table 6).

**Table 6:**
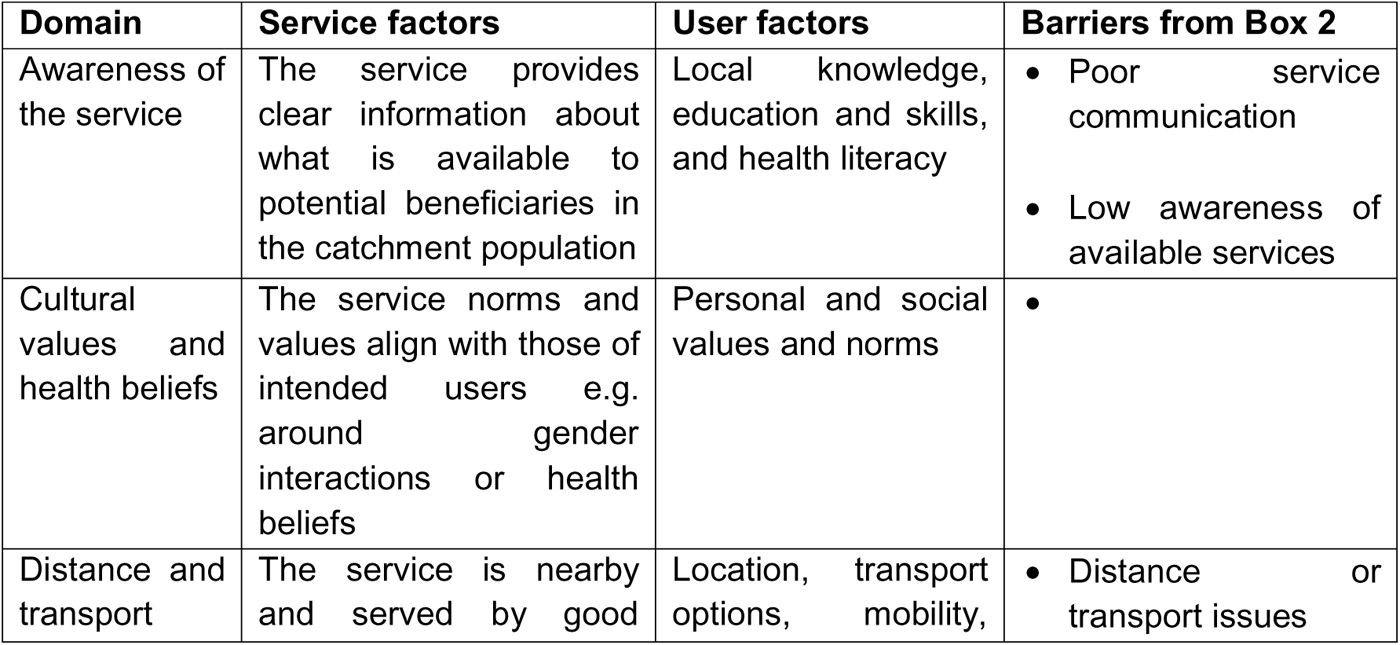
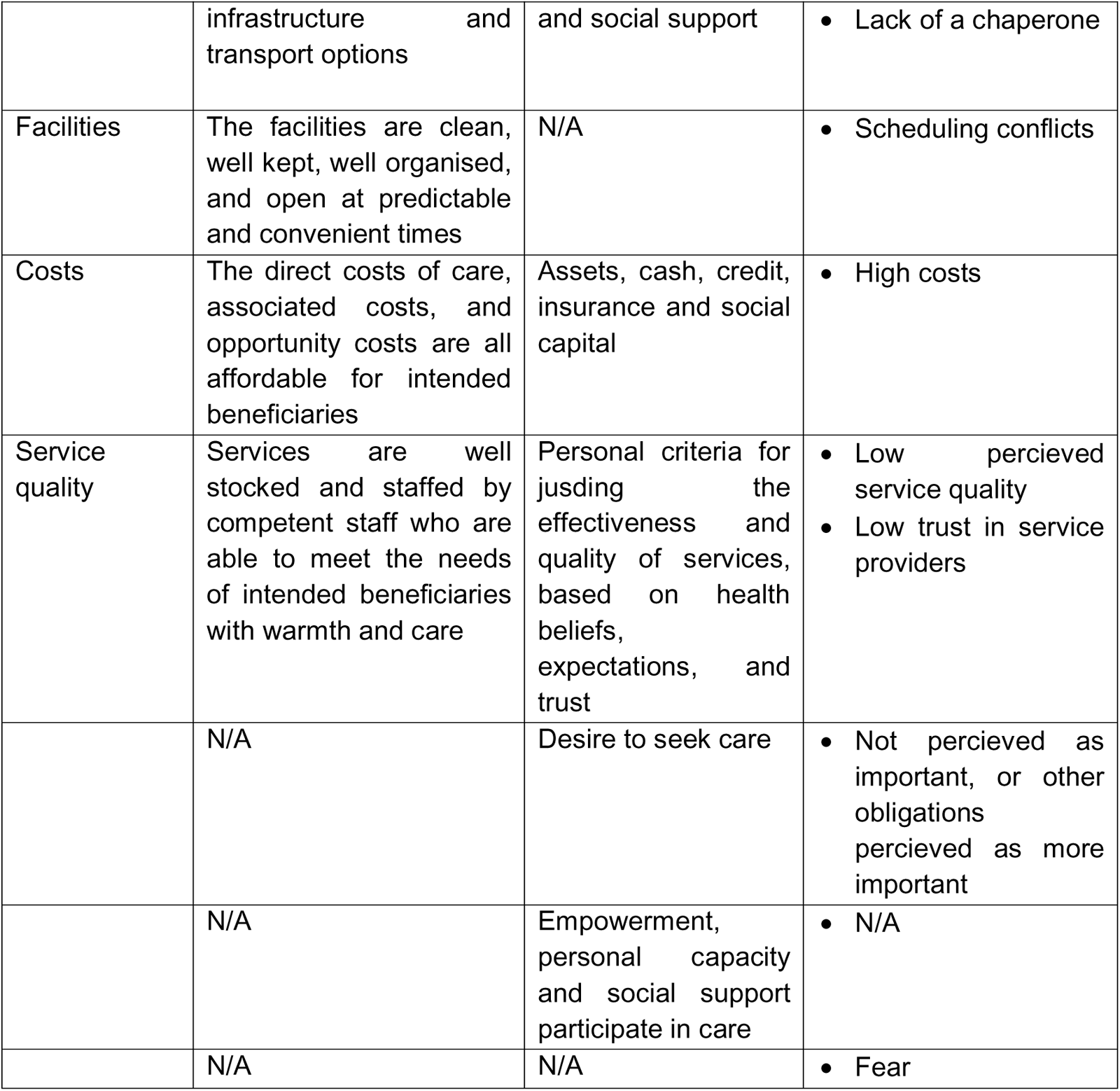
Mapping service and service user domains to common barriers from existing eye care research.

Finally, we reconfigured this table to create a deductive template that can be used to enter quotes during and directely after each interview. The whole point of using interviews rather than a (much cheaper and faster) survey is to be able to uncover barriers and potential solutions that the research team had not previously considered. As such, the template, interview prompts, and data collector training all emphasise the ‘other’ column.

**Figure 4:**
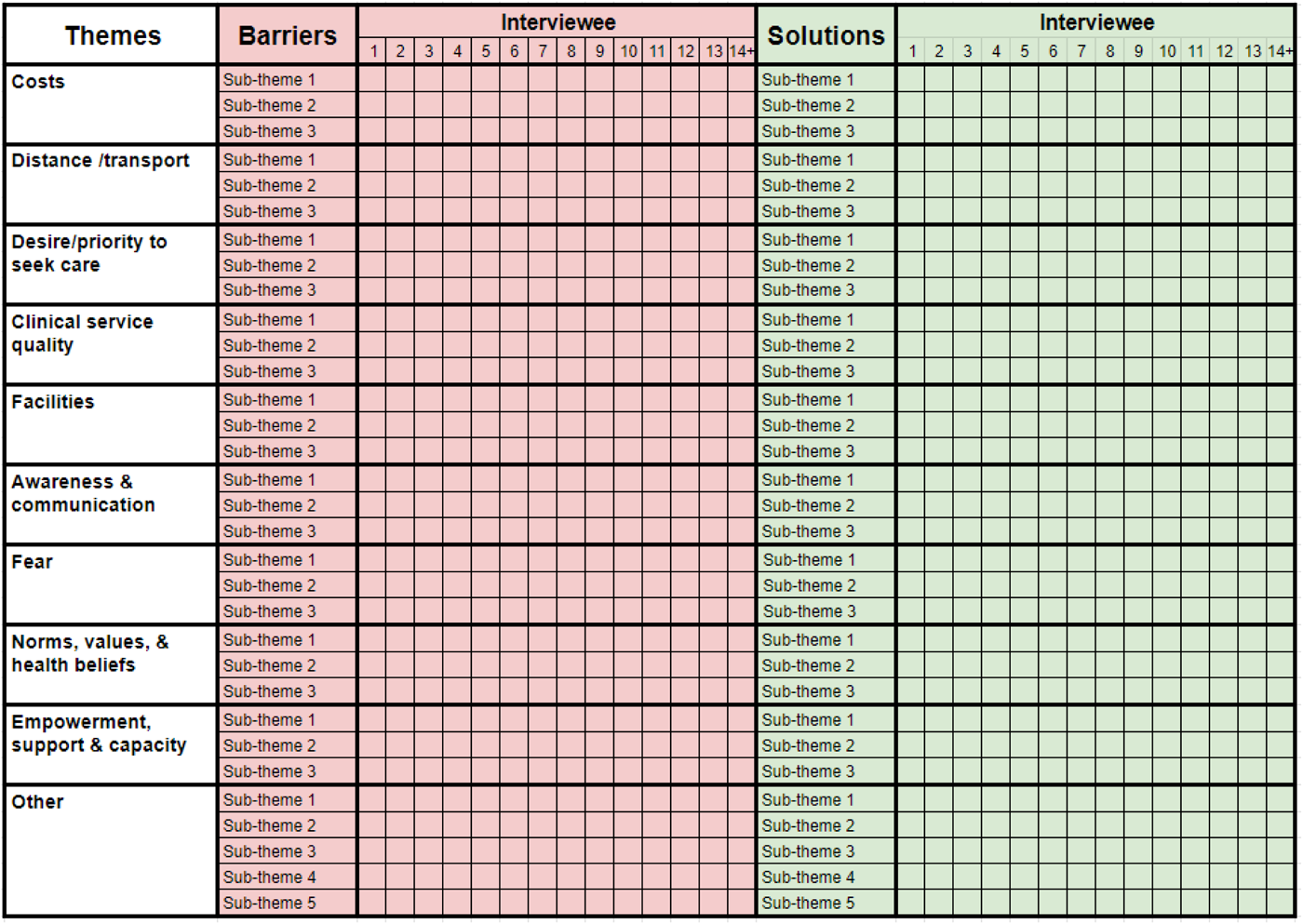
Our analytic matrix.

### Process for completing the matrix

During the interview, data collectors will expand the column width for the relevant interviewee number. They will type notes on each barrier into the relevant row, using the participant’s own words. Data collectors will repeat the process when asking for potential interventions that would have made it possible to attend, adding ideas to the matrix, supported by direct quotes. They will probe for further forms of service modification (which we are able to change) that would make a tangible difference.

Directly after the interview they will listen back to the audio recording to correct and expand upon quotes that they noted during the interview. All quotes will be directly translated into English. Data collectors will replace the ‘Sub-theme n’ text in the ‘Barriers’ and ‘Solutions’ columns with add their own (inductive) codes, for instance; ‘long queue at clinic’, ‘cost of spectacles’, or ‘rumours of rude staff’. The number of sub-themes is not limited; new rows can be added as required. As stated above, after a minimum of 14, interviews will continue until no new sub-themes emerge from two successive interviews. Data collectors will debrief with national research leads each day. The national research leads and the international research manager will collaboratively check quality and consistency of data entry, review all quotes and sub-themes, and assess when thematic saturation has been reached. Once qualitative data analysis is complete, all audio recordings will be deleted.

### Use of findings

Once saturation is reached, the wider research team will use the full matrix to generate a list of all the individual barriers and solutions that arose from the interviews. These may include things like sending SMS reminders, reducing the distance that people have to travel, or altering the way that people are counselled before being referred.

The long list of solutions will be reviewed by the programme funder and programme implementer to rule-out any service modifications that are completely unfeasible – such as paying people $100 to attend, or providing free individual transport for every participant. The short list of potential service modifications will form the basis of a survey that will be sent to a wider sample of non-attenders in order to identify the most promising actions at a generalisable level.

### Survey

As stated above, we will have a complete list of every non-attender belonging to the sociodemographic group with the lowest overall attendance rate. We will administer a telephone survey to a representative sample of non-attenders from this group, excluding all of those who have already been interviewed. We will use a 95% confidence interval, a 5% margin of error, and a conservative assumption that the total population size is 1 million people (with the same characteristics as the most marginalised group). This renders a sample size of 384.

We will use computer generated numbers to obtain a random sample of non-attenders to call. Data collectors will seek verbal audio recorded consent before reading through the full list of potential service modifications that arose from the interview stage. Respondents will be asked to rank each suggestion from 1-3 on a simple Likert scale:

1. It would make a big difference - i.e. if we introduced this change then you or people like you would definitely attend
2. It would make a moderate difference - i.e. it would greatly increase the chances, but it would not be enough by itself to guarantee attendance by itself
3. It would make a small difference - i.e. it might help a few people, but the impact is likely to be minimal

We will calculate the average score for each service modification and generate a ranked list. Workshop participants will review the ranked list and select the most promising service modification to implement and evaluate.

### Workshops

Our team already has formal agreements and pre-existing working relationships with Peek programme leads, programme funders, programme implementers, eye care policymakers, and community advisory boards in each location. In each country we will invite these stakeholders to a 60-90 minute workshop to review the study findings and select one or more service modifications to implement. Workshop discussion will be led in English (the working language of the project in each country) by a facilitator from our research team. The researcher will present a brief overview of the barriers and potential solutions suggested by non-attenders and their proxies, and then facilitate discussion to explore the groups’ perceptions of which barriers they can realistically address, and which solutions offer the best balance of impact (based on survey respondent scores), cost, risk, and feasibility. The aim is to identify promising service modifications that can be deployed and tested using RCTs to equitably improve access to care.

The process of decision-maker group discussion aligns with rapid methods that use group discussion with the ultimate research users as a key part of data analysis, interpretation, and application. The workshop will close with the identification of the most promising service modification to test and discussion of next steps.

### Output

The primary output of this mixed-methods study will be the selection of one or more feasible service modification(s) that has been identified by intended service users and agreed by service managers. This process will conclude during the workshops held in each country. The selected service modification(s) will be tested across the relevant programmes using an adaptive randomised trial design, as part of the broader ‘IM-SEEN’ approach.

## Ethical Considerations

### Institutional review

We will seek ethical approval from the LSHTM ethics committee and all relevant ethics committees in Botswana, India, Kenya and Nepal.

### Consent

#### Consent to be contacted for recruitment

In the screening stage that takes place before this project’s elicitation activities, written tick-box consent will be sought to use personal and contact data to recruit non-attenders for this current study. Our team is fully embedded in the screening programmes in each country, and there are memoranda of understanding in place that govern the sharing of data between parties.

> *Consent wording used at screening:*

> *I understand that my / my child’s anonymised data may be shared with other researchers or online in a public repository for research. I understand that I may be contacted by Ministry of Health partner organisations inviting me to participate in future studies to improve access to eye care services. I understand that I can call [phone number] for free to ask any questions; that my decision will not affect the care that I / I or my child receives; and that I can change my mind at any time*.

#### Consent for telephone interviews

For the qualitative interviews, we will call potential participants and provide information about the purpose and risks of the telephone interview using an appropriate version of the Botswana script shown below. Potential participants will have the opportunity to discuss the study and ask questions.

> *Hello, my name is ___. I am a researcher from the University of Botswana, working with the Ministries of Health and Basic Education on the Pono Yame eye screening programme.*

> *Your child recently had their eyes screened at school and was found to need further assessment. Our records indicate that, like many other children, they were unable to attend that appointment*.

> *You are being contacted because you have previously provided consent to be contacted by Ministry of Health partner organisations regarding research being conducted for eye care services. I am calling to invite you to participate in a 30-minute interview. Your participation is completely voluntary. This means that you do not have to do it unless you want to*.

> *We want to understand the barriers that prevented your child from attending. We are also asking about how we could change the Pono Yame programme to make it easier for children to attend appointments*.

> *Before agreeing, here is the background information that you need to know:*

> *We have invited you because, like many other referred children, your child did not attend. We want to hear about the issues that you personally faced that prevented your child from attending, and your ideas on how to make things easier. In total we are aiming to interview about 20 people*.

> *Who are we? I work with a group of researchers from the University of Botswana and the London School of Hygiene and Tropical Medicine. We are working to improve the national Pono Yame eye screening programme that will visit every school in the country. The leaders of the research are Prof Keneilwe Motlhatlhedi and Dr Luke Allen*.

> *We will take the responses from all of the interviews and discuss the ideas for improvement with the leaders of the national programme. We hope to use your suggestions to make the programme work better*.

> *We are also conducting a set of face-to-face interviews and online surveys with other parents and guardians. We want to compare the responses we get from these different approaches*.

> *In this 30-minute interview there are no risks to you or your child. If you agree to take part, we will send you a 100 pula airtime voucher to compensate you for your time. It is important to note that agreeing or declining to take part does not have any impact on your child, their schooling, or the services they receive*.

> *You can stop the interview at any time*.

> *I will record the interview. Our team will anonymise your data and keep it safe and secure on a password-protected computer in London. When the study is completed, we will write-up our findings and publish them online so that other researchers can use the information to help people in other places*.

> *The University of Botswana and London School of Hygiene and Tropical Medicine ethics committees have both approved this study*.

> *You can ask me any questions you like now. I can also give you the email address and phone number of the lead researchers if you’d like to contact them directly [provide the contact details for BK, Keneilwe or Luke as required]. If you have any other concerns I can also give you the contact details for the London School of Hygiene and Tropical Medicine Research Governance and Integrity Office*.

> *Do you have any questions?*

> *Are you happy to begin the interview?*

#### Consent for the telephone survey

For the telephone survey, we will call potential participants and provide information about the purpose and risks of the telephone interview using an appropriate version of the Kenyan script shown below. Potential participants will have the opportunity to discuss the study and ask questions.

> *Good morning/afternoon*

> *My name is …. and I’m calling from the Vision Impact Project eye screening programme. We saw you a few weeks ago and referred you to the local clinic, but we did not see you on your appointed day*.

> *In fact, half of all people who were referred did not attend. We have sought feedback on ways we could improve our service, and I wanted to ask you which of the ideas we have stand the best chance of helping people like you to access care. It should take approximately 15 minutes of your time*.

> *If you are happy to proceed, I need to tell you a bit more about the survey. I will then double-check that you are still happy to proceed*.

> *I will ask you about a set of potential changes that we are thinking about making. I will ask you to rate each one in terms of how likely you think it is to make a difference at helping people access our clinics*.

> *Your responses will help us to shape and improve our services for others, but there are no direct benefits to you for taking part. Thinking about the issues that prevented you from getting care may be distressing to you. If you face any discomfort because of the questions asked, you can skip any question or ask to end the call whenever you choose*.

> *If you don’t want to take part, that’s ok. You can drop out of the survey at any point. Your decision will not affect your health care or your future relations with the Vision Impact Project in any way*.

> *Your anonymised answers will be combined with those from other people and kept safe and secure on password-protected computers in Nairobi and London. None of the data will be used for commercial use. We will publish our findings in a research journal and in a public repository so that other researchers can learn from what we find. You personal information will not be included in our findings and there is no way that you can be identified from any of the reports that we will produce*.

> *If you have any questions, you can ask me now, or I can put you in contact with the study coordinator - Sarah Karanja from Kenya Medical Research Centre. If you have any questions about your rights as a research participant, I can connect you with the Kenya Medical Research Centre Ethics team who approved this survey*.

> *Does that all make sense? Do you have any questions for me?*

> *Are you happy for me to start?*

#### Consent for participation in the workshop

All participants will be participating in the workshop as a routine part of their duties in connection with the respective eye programme. As such, consent is not required. The only output from this workshop will be the intervention(s) that will be implemented and evaluated using RCTs.

##### Risks and strategies to mitigate

The risks to participants from the interviews, survey, and focus group discussion are low and there are no physical risks. Dwelling on the issues that prevented attendance may cause psychological distress. Data collectors will be trained to supportively manage mild levels of distress and will signpost participants to other sources of support if participants become moderately or severely distressed.

Any issues, complaints or concerns will be reported to the principal investigators. Participants will be provided with their email addresses and office phone numbers. Participants will also be given the number of the local field coordinator for operational queries, and the LSHTM RGIO contact details for any other concerns about the conduct of the study.

We will compensate telephone interviewees for their time with an airtime voucher worth 100 BWP / 500 KES / 800 NPR (approximately £5). The voucher will be sent via SMS to telephone interviewees. Given the lower time and cognitive burden, survey responders will not be offered reimbursement, neither will workshop participants as quality improvement is a core part of their role.

All data collected will be encrypted and stored on secure servers protected with strong authentication controls including two-factor authentication. All data will be processed and safeguarded in compliance with the EU and UK’s General Data Protection Regulation (GDPR). Data will be anonymised and kept confidential. After 7 years all study data will be destroyed. We have developed a robust Data Management Plan (Appendix 1).

## Discussion

The series of elicitation elements in this study will produce a list of barriers to accessing eye health services, as perceived by patients or their proxies, as well as insight into what service modifications may be most useful for overcoming these barriers. The survey and workshop will refine this list, identifying those service modifications that are deemed to be most impactful by a representative sample of non-attenders, as well as offering the optimal balance of impact, cost, and risk by programme managers.

Whilst our analytic framework is grounded in the literature, the obviation of transcription and dual coding by highly trained qualitative researchers clearly limits the reliability of the interview findings. We have deliberately sought to develop a method that can be deployed in low-resource settings where there are not necessarily qualitative researchers available and time is at a premium. Previous work has shown that rapid qualitative methods led by less-experienced research assistants are able to generate valid findings when the subject matter is not overly complex. Seeking a list of potential barriers and solutions meets these criteria.

The highest-ranked potential service modifications will be presented to local and regional policymakers and stakeholders to garner their views on which should be prioritised for implementation, based on their likely impact, feasibility, cost, and potential risks. Stakeholders include community advisory board representatives in each setting. By having community members assist with analysis and interpretation of study findings, this design provides a participatory approach to the selection of interventions and health service modifications that will be tested in subsequent work. Those responsible for funding and implementing the modifications will also play a role in reviewing data and selecting the most appropriate interventions to test.

Improvements in access to health services and health equity are the key component of this study as we seek to focus on the needs of the most marginalised groups of non-attenders. We aim to refine and apply these methods to address other areas blighted by inequitable and low access.

### Limitations

Despite the fact that phone penetration is high in the countries we are working in, not everyone has their own phone and it is also likely that members of the most disadvantaged groups will be the least likely to respond to our telephone interviews and surveys, as well as being the least likely to attend services. It is possible that those with access to phones have different opinions on barriers and interventions and this could bias the results. In terms of alternatives, postal surveys are problematic for a range of other reasons including the lack of addresses, poor reliability of the postal service, and issues with loss of data. In-person surveys would be the most robust way of ensuring that every voice is heard, but we do not have the time or resources given the national scale of the programmes.

### Dissemination

Our findings will be shared with lay representatives, community advisory board members, local and national programme funders and implementing partners, Peek Vision, and national eye care policymakers. No participant names or identifiable information will be used. The study findings will also be disseminated during quarterly review meetings with implementing partners, community workers and representatives from the county health management committee, and bi-annual partner meetings. We will also present our findings at national, regional and/or international conferences.

## Data Availability

All data produced in the present work are contained in the manuscript.

## Study status

At the time of manuscript submission we have piloted the approach in Kenya and obtained ethical approval for every country. We have not started recruitment or data collection in Botswana, India, or Nepal.

## Study coordination centre

London School of Hygiene & Tropical Medicine. This trial will adhere to the principles outlined in the International Conference on Harmonisation Good Clinical Practice (ICH GCP) guidelines, protocol, and all applicable local regulations.

## Patient and public involvement

Lay representatives from our community advisory boards in Botswana, India, Kenya and Nepal reviewed the original draft, provided feedback on the proposed methods, and reviewed the final version of this protocol, as well as contributing to earlier related studies as part of the broader IM-SEEN project.

## Acknowledgements

Thanks to our community advisory board members who reviewed this protocol and continue to support the IM-SEEN project: Mr. Niranjan Prasad Yadav in Nepal; Mr Ashok Saxsena in India; Theresa Kowa and Oratile Moatlhodi in Botswana; and Paul Ringera in Kenya.

## Appendix: Data Management Plan

## 1. DATA SOURCES AND DATA COLLECTION PROCESSES

The research objectives require the collection of quantitative survey data, as well as qualitative data in the form of audio recordings and quotes from study participants. Table 1 below outlines the data fields to be collected throughout the various stages of the data collection process. All data will be treated as personal data for the purpose of data capturing and processing, as collectively, it can be combined in a way that could make it identifiable.

Data from the initial screening process will be collected in Peek powered Eye Health School and Community Programmes using Peek’s Capture application. During the initial screening process only basic and non-personal identifying data is collected, with the exception of telephone number. Following initial screening, all those identified as requiring referral will be asked to provide sociodemographic data to enable us to monitor the equity performance of our programmes e.g. are certain ethnic groups more likely to be screened? The additional sociodemographic indicators are outlined in table 1 below. Based on the visual acuity threshold set prior to screening, the Peek Capture automatically informs the data collector whether the attendee may potentially need onward treatment. For those screened negative no further data is collected. Only for those screened positive is further information collected. This ensures data collection is kept to an absolute minimum maintaining privacy and ensuring compliance with data protection regulations. For those screened positive, additional information is collected, but the data is always minimised to ensure only the required data is collected at each stage of the service.

Following triage of individuals who had screened positive, a four-stage rapid exploratory sequential mixed-methods study design will be used to evaluate barriers to health access among non-attenders who had been flagged for onward treatment. Telephone interviews will be conducted among 60 non-attenders, purposively selected from socio-demographic groups with the lowest overall attendance rates. The aim of the telephone interviews is to explore and evaluate their perceived barriers to clinic attendance, and develop a list of potential solutions. Once interventions and service modifications have been identified, these will be tested through a series of pragmatic, embedded, adaptive parallel, multi-arm randomized control trials (APT). The intention of the APT is to continuously improve attendance rates, particularly amongst those groups with the lowest engagement rates overall. Table 1 outlines each of the data collection phases, the data fields to be collected, and the study populations of each of the stages discussed.

**Table 1:**
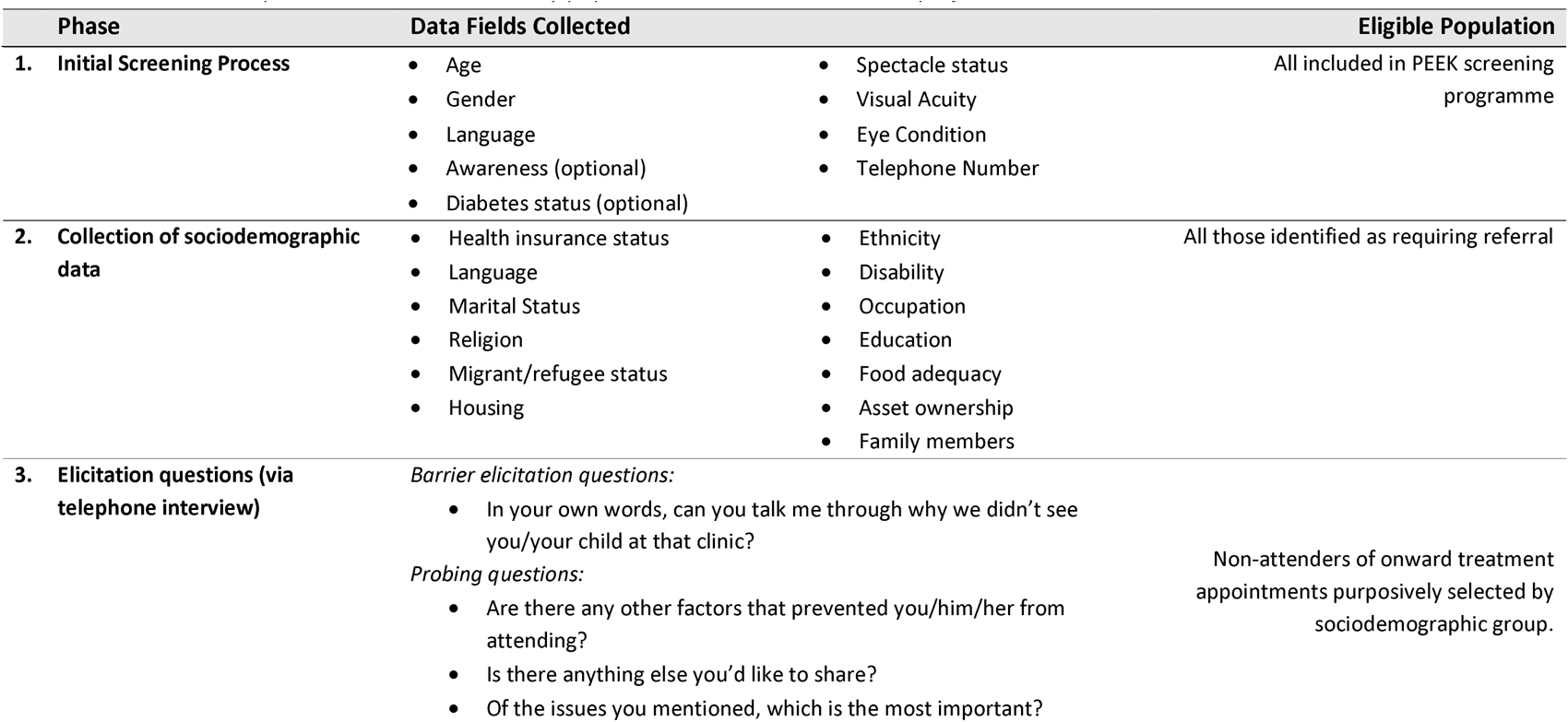
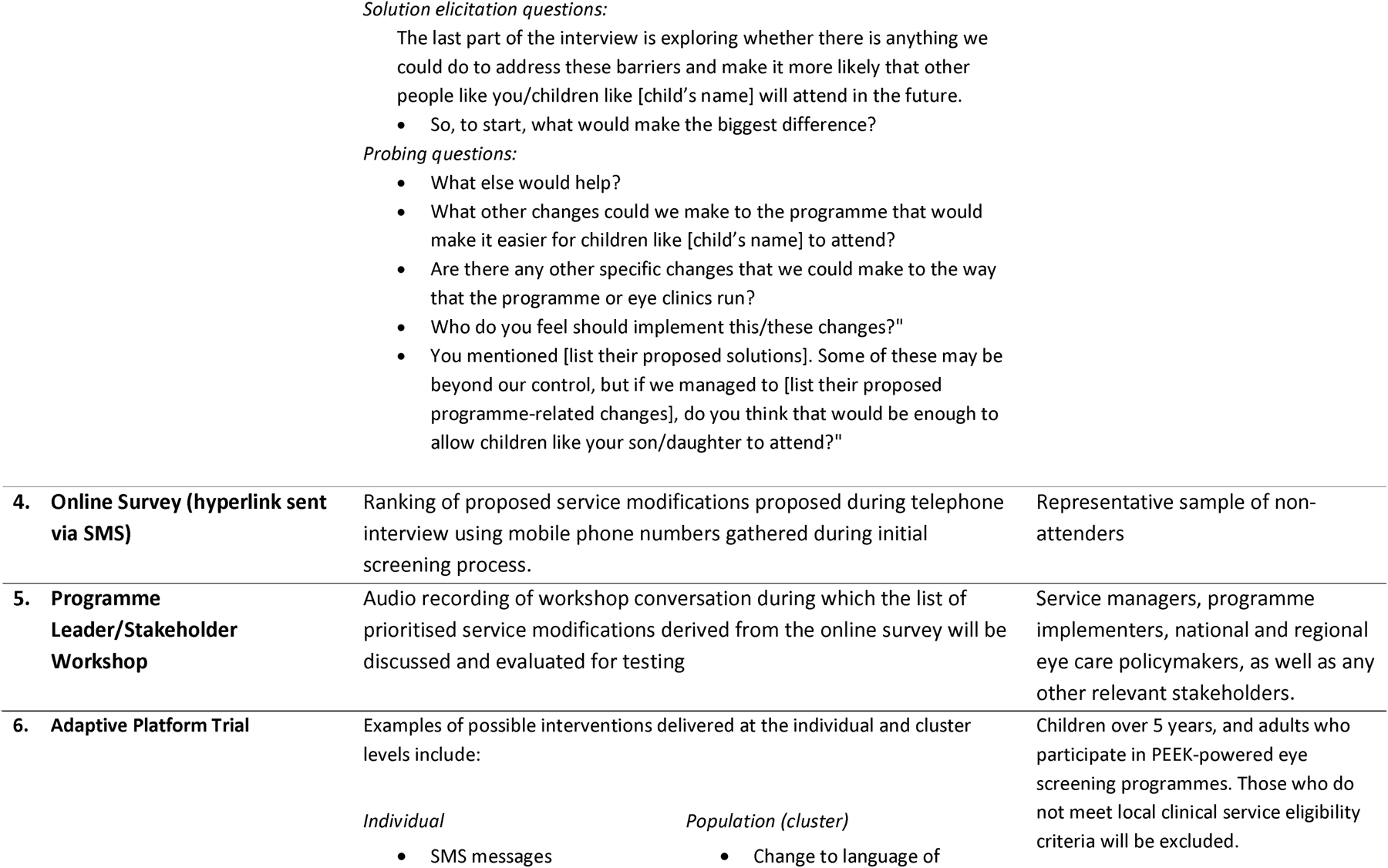
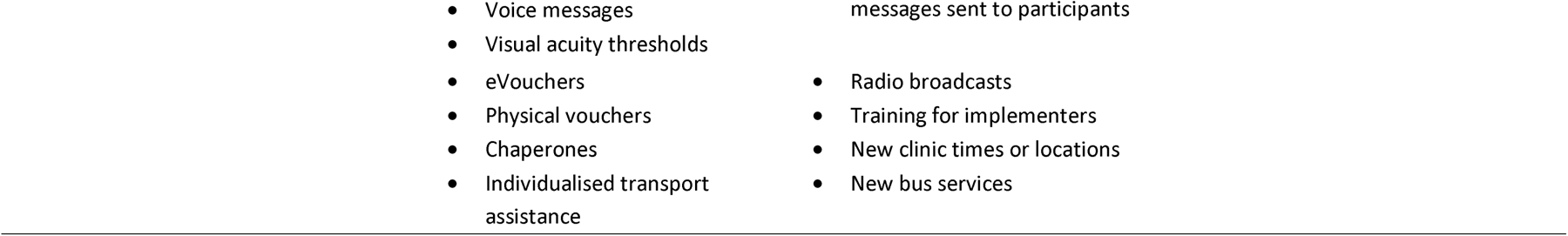
Data collection phases, data fields and study populations for broader I’M SEEN project.

## 2. DATA COLLECTION TOOLS

<u>Various data collection tools will be used to populate the data fields outlined in</u> table 1.

<u>Quantitative Data:</u>

- <u>Android Mobile Devices</u> – Survey data, and data derived from the APT (phases 1,2, 4 and 6) will be collected by Peek’s implementing partners using Android devices through the Peek Capture application. Peek Capture enforces security controls that include strong device passcodes and native Android encryption. Data stored is time limited, the device syncs via an encrypted connection with a Peek managed server, the data is then deleted to minimise the risk of data stored on the device. The APT will be embedded within Peek software used in parallel with a Bayesian algorithm that will be used to autonomously run response adaptive trials.

<u>Qualitative Data:</u>

- Play Verto – The online survey will be administered through Play Verto, a play-based online survey group who have worked with the United Nations and others to develop engaging short surveys that have impressively high response rates in low- and middle-income countries. The survey will be sent as a hyperlink in an SMS. PlayVerto will gather, store and process. After, they will transfer (anonymised data) it to LSHTM who will perform further processing and storage. LSHTM will share aggregate anonymised findings with partners and in public domain.
- Data Abstraction Matrix: During the telephone interviews, data collectors will directly enter notes, quotes, open codes, and abstractions into a matrix. Data gathered, processed and stored by local partner organization. Then shared with LSHTM (fully-anonymised responses to be shared).
- Audio Recordings – Telephone interviews will involve verbal communication and discussion, and thus will be collected and stored using digital audio-recording methods.

**Software:**

- Peek Capture - is an application that runs on Android devices that supports eye health screening and referral pathways to treatment
- Peek Admin - is a web based data platform application that is used to view the data collected by Peek Capture, it tracks the Programme progress, provides insights and helps ensure no one is left behind.
- Play Verto – is a play-based online survey group who have worked with the United Nations and others to develop engaging short surveys that have impressively high response rates in low- and middle-income countries.
- STATA and R, and Excel will be used to analyse the data exported from Peek Admin

**Hardware:**

- Peek servers are hosted on Amazon Elastic Compute cloud-based virtual machines running Amazon Linux.
- Android devices, locally managed by Peek’s implementing partners.

## 3. DATA-RELATED ACTIVITIES

| Task | Description |
| --- | --- |
| Start gathering SES data | In month 1 we will start gathering sociodemographic data from:<br>• a representative sample of all those presenting to be screened<br>• all those identified with an eye care needs and referred on for treatment<br>These data will be transferred from Android devices in the field to Peek Admin, hosted on AWS.<br>Note that Peek programmes run continuously and we intend to gather data from participants in every programme so that we can promote equitable service delivery. |
| Clean SES data | Routine manual data cleaning will be conducted periodically by Peek administrators. Internal software guardrails will pick up simple errors |
| Analyse SES data | Every month we will perform simple descriptive statistical analysis of presentation rates and treatment attendance rates by SES category.<br>The output of this analysis will be anonymised and presented as mean attendance rates for each SES subgroup e.g. males x%, females z%. |
| Conduct telephone interviews, online surveys and stakeholder workshop | In order to better understand barriers to accessing eye services a series of activities will be conducted through a four-stage sequential mixed-methods approach. These include:<br>1. Telephone Interviews – Telephone interviews will be conducted with non-attenders, purposively selected from subgroups with the lowest attendance rates.<br>2. Following telephone interviews, a single list of suggested solutions will be compiled<br>3. Online survey – An online survey will be conducted among a representative sample of non-attenders to rank mooted interventions/service modifications.<br>4. Stakeholder workshop – Programme leaders and key stakeholders will then select one or more of the highest ranked interventions to test, based on impact, feasibility, risk and cost. |
|  | Following completion of this process, data will be analysed to elicit barriers to care and recommended interventions/service modifications to improve attendance rates. |
| Testing of service modifications through APT | An automated adaptive platform trial (APT) will iteratively test a series of interventions selected with intended service beneficiaries to increase attendance rates among marginalised groups. This will be done through a Bayesian, embedded, pragmatic, superiority, adaptive platform trial platform that will use response adaptive randomisation. |

### Quality checks

- Errors are flagged at the point of data entry by software that only accepts pre-specified responses e.g. phone numbers must be comprised of a set string length of digits.
- The software has built-in logic steps
- We will institute training and supervision for all data collectors
- Application logging, audit trails and alerting direct administrators to given issues post-collection e.g. when SMS messages fail to be delivered
- Post-collection human data checking using the Peek Admin programme e.g. for ID disambiguation

5. How will you address ethical & legal issues within your research?

- What permissions are needed? E.g. to collect data in country, analyse data for specific purpose, share data
- From whom must approval be obtained? E.g. study participant, ethics committees, data provider
- How will permissions be provided? E.g. ask participants to sign a consent form, sign a Data Transfer Agreement

## 4 PERMISSIONS

Local permissions for Peek powered eye health programmes are already in place. This is in the form of data processing agreements with Peek and the local MoH and/or local implementing partner. This provides a legal agreement between the parties that the data can be collected and processed. The proposed research will be authorised by the same parties to ensure full transparency and the data collection and processing will be managed under the same data processing agreement.

We will obtain written informed consent to collect, analyse, and publish anonymised aggregate participant data in peer-reviewed journals and online open-access data repositories. Individuals will not be identifiable.

In line with UK guidance on risk-adapted approaches to obtaining informed consent, participants will provide consent by ticking a box underneath the following statement:

> *“I understand that my anonymous data may be shared with other researchers or online, and that I will not be identifiable from this information. I understand that my decision will not affect the care that I receive, and I am free to change my mind anytime I like.”*

Consent will be obtained when participants initially present for screening.

For screening programmes that include children (<18 years), we will seek consent from their parents/legal guardians using the following statement, sent home on a paper form along with the generic participant information leaflets before screeners visit the school:

> “I *understand* that my child’s anonymous data may be shared with other researchers. I understand that my child will not be identifiable from this information. I understand that my decision will not affect the care that my child receives, and I am free to change my mind anytime I like.”

Approval will be sought from research ethics committees at LSHTM and each of the countries where screening takes place.

## 5 DOCUMENTATION

Standard operating procedures and an overall study protocol will be developed in line with LSHTM research guidance to cover all aspects of the research project.

Standardised online training modules have been delivered for programme implementing partners tasked with data collection in the field.

Training will be delivered to all project staff to ensure that they understand the requirements and are able to follow the SOPs.

We have a data compendium which describes the custom sociodemographic variables that we will collect in each country,

## 6 DATA STORAGE AND SECURITY

Data collection, management and storage for this study will be managed by seven entities described below:

A. Peek Vision Capture Application
B. Play Verto
C. The London School of Hygiene and Tropical Medicine
D. Botswana: The University of Botswana
E. India: Dr Shroff Charity Eye Hospital
F. Kenya: Kenya Medical Research Institute?
G. Nepal: Nepal Netra Jyoti Sangh

### Peek Capture Application

#### Pre research data collection and storage in Peek powered eye health programmes

The data will be collected in Peek powered Eye Health School and Community Programmes using Peek’s Capture application. Data will be collected by Peek’s implementing partners using Android devices through the Peek Capture application. Peek Capture enforces security controls that include strong device passcodes and native Android encryption. Data stored is time limited, the device syncs via an encrypted connection with a Peek managed server, the data is then deleted to minimise the risk of data stored on the device. h

The data is stored on a Peek managed server hosted in a Virtual Private Cloud (VPC) utilising the Amazon Web Services (AWS) Cloud. Each Peek powered programme is hosted on it’s own dedicated server and a VPC that will reside in the UK/EU ensuring all of the data privacy safeguards as governed under the GDPR. All data collected is securely stored in AWS data centers which are state of the art, utilising innovative architectural and engineering approaches. More information, including a virtual tour, can be found by visiting the link <u>here</u>.

Throughout the eye health programme life cycle only approved implementation partners and Peek team members have access to programme data. Access is strictly controlled through the Peek Admin web based data platform application. This is used to view the data collected by Peek Capture, it tracks the Programme progress, provides insights and helps ensure no one is left behind.

**Peek Capture security:**

- Peek Capture is installed on implementing partners managed Android devices
- Peek Capture enforces security controls that include strong device passcodes and native Android encryption.
- Data stored is time limited, the device syncs via an encrypted connection with a Peek managed server, the data is then deleted to minimise the risk of data stored on the device.

**Peek Admin security:**

- Strong passwords, minimum of 12 characters, password strength meter where only ‘strong’ is accepted, blacklist passwords are enforced to ensure easily guessed and passwords found in data breaches cannot be used.
- 2-Factor Authentication to protect user account security.
- User access permissions are controlled through account privileges, this controls scope of programme so access is restricted and limited to only what a user requires for their work, admin privileges are restricted to only those that require the access, account management and patient level reporting.
- Accounts disable automatically after 60 days of inactivity.
- User access reviews available for implementing partners to ensure leavers and inactive accounts are removed.

**Peek Platform Data Security Assurance:**

Peek is an International Standardisation Organisation (ISO) 27001 certified organisation. ISO 27001 certification requires an annual audit by an accredited external auditing body who verify compliance with the industry best practice information security controls.

Peek servers hosted in a Virtual Private Cloud (VPC) utilising the Amazon Web Services (AWS) Cloud. Each Peek powered programme is hosted on it’s own dedicated server and a VPC that will reside in the UK/EU ensuring all of the data privacy safeguards as governed under the GDPR. All data collected is securely stored in AWS data centers which are state of the art, utilising innovative architectural and engineering approaches.

More information, including a virtual tour, can be found by visiting the link below: https://aws.amazon.com/compliance/data-center/.

Annual penetration tests conducted by a 3rd party specialist security testing company. The purpose of the test is to verify whether robust security mechanisms are in place to prevent unauthorised users from accessing data and infrastructure. This penetration test includes:

- Identification of potential vulnerabilities occurring in the application and defining possible attack scenarios conducted with techniques typical for attacks on web applications;
- Simulated attacks from the perspective of an anonymous and standard user;
- Testing API endpoints from the perspective of an anonymous and standard user, including mechanisms such as user authentication, access control, and data validation;
- Security assessment of our infrastructure against the latest industry standard AWS CIS Foundations Benchmark.

The AWS Compliance Program provides further assurance and understanding of the robust controls in place to maintain security and compliance in the cloud. AWS regularly achieves third-party validation for thousands of global compliance requirements that are continuously monitored to meet security and compliance standards for the most sensitive data and privacy requirements. AWS supports more security standards and compliance certifications than any other offering, including PCI-DSS, HIPAA/HITECH, FedRAMP, GDPR, FIPS 140-2, and NIST 800-171, helping satisfy compliance requirements for virtually every regulatory agency around the globe. More information can be found by visiting https://aws.amazon.com/compliance/programs/.

**Peek Platform Data Security Controls:**

*Peek Servers:*

Peek servers hosted in a Virtual Private Cloud (VPC) utilising the Amazon Web Services (AWS) Cloud. Each Peek powered programme is hosted on it’s own dedicated server and a VPC that will reside in the UK/EU ensuring all of the data privacy safeguards as governed under the GDPR.

Server OS is Amazon Linux ustlising AWS AMIS to provide base images for our system drives and enhances security by focusing on two main security goals, limiting access and reducing software vulnerabilities. Security updates are applied automatically to test once a week and then rolled out a week later automatically to other environments

*Docker:*

Peek server software runs in Docker containers. Docker shields application software from variations in platform and co-hosted software. It ensures that development, test and production environments run the same context as one another to ensure consistent, predictable behaviour. Peek servers also use docker swarm mode to achieve failsafe reliability and replication of Mongo databases.

*Databases:*

Server data is stored in Mongo databases, a fast, scalable, json document database. Peek infrastructure uses a Mongo replica set across two hosts. There are two replicas each holding a full copy of the data and one arbiter. The arbiter is only used for the election of a new master if one of the nodes was to become unavailable. The Mongo database and journal are held on AWS Secure EBS volumes. This provides 256-bit AES encrypted using a key managed under the Amazon Key Management Service.

Amazon Key Management Service, allows us to create and manage cryptographic keys and securely control their use across a wide range of AWS services and within our applications. AWS KMS is a secure and resilient service that uses hardware security modules that have been validated under FIPS 140-2 to protect the encryption keys. AWS KMS also integrates with AWS CloudTrail providing us with secure logs of all key usage. Backups on S3 are also encrypted using keys managed by AWS Key Management Service.

*Logging and Monitoring:*

Peek Server and Mongo Server logs and uploaded to AWS Cloudwatch for storage and monitoring. AWS Cloudwatch collects monitoring and operational data in the form of logs, metrics, and events and alerts us immediately of problems in any environment, both application and infrastructure.

*Network Security:*

AWS Security groups are used to provide firewall-like network access control and allow inbound traffic on HTTP and HTTPS ports. Outbound traffic is permitted on any port. The SSH traffic is restricted to subnets associated with devops engineers and the deployment servers. TLS 1.2 is used to secure traffic between device or browser and server.

Operational access to the AWS console is protected with AWS IAM MFA which uses 2-Factor Authentication and ensures that access to AWS is restricted to users with knowledge of password and possession of a specific approved mobile device. Automated access to the AWS API uses AWS Roles with restricted privileges needed for housekeeping, logging and alarm maintenance. No user use is made of Access Keys to eliminate the vulnerabilities of file-system-based credentials.

*Threat Detection:*

AWS Guard Duty is enabled, this provides a threat detection service that continuously monitors for malicious activity and unauthorised behaviour to protect access, workloads and data. The service utilises up-to-date threat intelligence feeds from AWS, CrowdStrike, and Proofpoint and continuously evolves through machine learning.

*Backups:*

An Image is maintained of the Server Host using AWS AMI to ensure continuous availability.

A snapshot of the encrypted data volume, containing database and journal, is taken four times daily. Snapshots are retained for two weeks. Access to the snapshots is strictly controlled. Old backups are automatically deleted after 90 days. Backups are stored on AWS S3 storage, also encrypted providing 256-bit AES encryption. The backups are stored across AWS multiple availability zones, this ensures that the data resides in multiple data centres separated geographically and stored in AWS secure data centres.

Additionally, a further backup is made off AWS. Off-AWS backups are replicated to Google Cloud daily via Google Transfer service to identically named buckets and files with a retention policy of 90 days.

*Data Centres:*

All data collected is securely stored in AWS data centers which are state of the art, utilising innovative architectural and engineering approaches.

*Disaster Recovery:*

A full disaster recovery test is performed at least annually to ensure servers, applications and databases can be fully recovered within 24 hours.

### Play Verto

#### Play Verto Data capture tool

Data collection via our web-based application is all stored on a AWS RDS dedicated server, located in Ireland. This database utilises AWSs own encryption, AES-256 at rest, for maximum security. All data collected is securely stored in AWS data centers which are state of the art, utilising innovative architectural and engineering approaches. More information, including a virtual tour, can be found by visiting the link <u>here</u>.

Only approved team members have access to the data. Access is strictly controlled through the Play Verto’s Admin and AWS Admin. Where Password protection is required and the use of 2-factor authentication where applicable.

*Play Verto Capture security:*

- Play Verto is a web-based application therefore can only be accessed via a public URL.
- Play Verto enforces security controls that include strong device passcodes and 2-factor authentication where applicable…
- Data stored is encrypted via AES-256 encryption

*Play Verto Admin security:*

- We have a strong password policy in place for all our accounts, requiring a minimum length of 8 characters.
- 2-Factor Authentication to protect user account security.
- User access permissions are controlled through account privileges. So access is restricted and limited to only what a user requires for their work.

*Play Verto Platform Data Security Assurance:*

Play Verto complies with CyberEssentials Certification and IASME Governance Standard. Data collection via our web-based application is all stored on a AWS RDS dedicated server, located in Ireland. This database utilises AWSs own encryption, AES-256 at rest.

Monthly automated penetration tests conducted by <u>Detectify</u> The purpose of the test is to verify whether robust security mechanisms are in place to prevent unauthorised users from accessing data and infrastructure. We have maintain Threat score of 0 and 10/10, OSWASP SCORE (The worldwide non-profit organization Open Web Application Security Project (OWASP)’s list of the ten most common vulnerabilities, known as OWASP Top 10, is often used as a security standard. Detectify covers OWASP Top 10 and provides an easy way for you to see which categories you pass or fail.)

The AWS Compliance Program provides further assurance and understanding of the robust controls in place to maintain security and compliance in the cloud. AWS regularly achieves third-party validation for thousands of global compliance requirements that are continuously monitored to meet security and compliance standards for the most sensitive data and privacy requirements. AWS supports more security standards and compliance certifications than any other offering, including PCI-DSS, HIPAA/HITECH, FedRAMP, GDPR, FIPS 140-2, and NIST 800-171, helping satisfy compliance requirements for virtually every regulatory agency around the globe. More information can be found by visiting https://aws.amazon.com/compliance/programs/.

***Play Verto Platform Data Security Controls:***

*Play Verto Servers:*

Data collection via our web-based application is all stored on a AWS RDS dedicated server, located in Ireland. This database utilises AWSs own encryption, AES-256 at rest, for maximum security. Ensuring all of the data privacy safeguards as governed under the GDPR.

*Databases:*

Server data is stored in Mongo databases, a fast, scalable, json document database. Play Verto infrastructure uses a Mongo replica set across two hosts. There are two replicas each holding a full copy of the data and one arbiter. The arbiter is only used for the election of a new master if one of the nodes was to become unavailable. The Mongo database and journal are held on AWS Secure EBS volumes. This provides 256-bit AES encrypted using a key managed under the Amazon Key Management Service.

Amazon Key Management Service, allows us to create and manage cryptographic keys and securely control their use across a wide range of AWS services and within our applications. AWS KMS is a secure and resilient service that uses hardware security modules that have been validated under FIPS 140-2 to protect the encryption keys. AWS KMS also integrates with AWS CloudTrail providing us with secure logs of all key usage. Backups on S3 are also encrypted using keys managed by AWS Key Management Service.

*Logging and Monitoring:*

Play Verto Server and Mongo Server logs and uploaded to AWS Cloudwatch for storage and monitoring. AWS Cloudwatch collects monitoring and operational data in the form of logs, metrics, and events and alerts us immediately of problems in any environment, both application and infrastructure.

*Network Security:*

AWS Security groups are used to provide firewall-like network access control and allow inbound traffic on HTTP and HTTPS ports. Outbound traffic is permitted on any port. The SSH traffic is restricted to subnets associated with devops engineers and the deployment servers. TLS 1.2 is used to secure traffic between device or browser and server.

Operational access to the AWS console is protected with AWS IAM MFA which uses 2-Factor Authentication and ensures that access to AWS is restricted to users with knowledge of password and possession of a specific approved mobile device. Automated access to the AWS API uses AWS Roles with restricted privileges needed for housekeeping, logging and alarm maintenance. No user use is made of Access Keys to eliminate the vulnerabilities of file-system-based credentials.

*Threat Detection:*

AWS Guard Duty is enabled, this provides a threat detection service that continuously monitors for malicious activity and unauthorised behaviour to protect access, workloads and data. The service utilises up-to-date threat intelligence feeds from AWS, CrowdStrike, and Proofpoint and continuously evolves through machine learning.

*Backups:*

An Image is maintained of the Server Host using AWS AMI to ensure continuous availability.

A snapshot of the encrypted data volume, containing database and journal, is taken four times daily. Snapshots are retained for two weeks. Access to the snapshots is strictly controlled. Old backups are automatically deleted after 90 days. Backups are stored on AWS S3 storage, also encrypted providing 256-bit AES encryption. The backups are stored across AWS multiple availability zones, this ensures that the data resides in multiple data centres separated geographically and stored in AWS secure data centres.

Additionally, a further backup is made off AWS. Off-AWS backups are replicated to Google Cloud daily via Google Transfer service to identically named buckets and files with a retention policy of 90 days.

*Data Centres:*

All data collected is securely stored in AWS data centres which are state of the art, utilising innovative architectural and engineering approaches.

*Disaster Recovery:*

A full disaster recovery test is performed at least annually to ensure servers, applications and databases can be fully recovered within 24 hours.

**EXPORT DATA SHARING FOR ANALYSIS** At the analysis stage pseudo-anonymised data will be exported in an encrypted zip file CSV file to LSHTM researchers to perform statistical testing. The zip file will be saved on the protected LSHTM server and only named project staff will be given access. Passwords will be sent separately. We will only ever export the minimum data required for the analyses.

**Labelling conventions**

1. Keep file names short, meaningful and easily understandable to others.
2. Order the elements in a file name in the most appropriate way to retrieve the record.
3. Avoid unnecessary repetition and redundancy in file names and paths
4. Avoid obscure abbreviations and acronyms. Use agreed University abbreviations and codes where relevant.
5. Avoid vague, unhelpful terms such as “miscellaneous” or “general” or “my files”
6. Use capital letters to delimit words, as the preferred option, although underscores (_) or hyphens (-) may add clarity, they make the file name longer.
7. For numbers 0-9, always use a minimum of two digit numbers to ensure correct numerical order (e.g. 01, 02, 03 etc.)
8. Dates should always follow same format: YYYY-MM-DD e.g. 2017-04-25
9. When including a personal name give the family name first followed by initials, with no comma in between e.g. SmithAB
10. Avoid using common words such as ‘draft’ or ‘letter’ at the start of file names unless doing so will make it easier to retrieve the record.
11. Use alphanumeric characters i.e. letters (A-Z) and numbers (0-9). Avoid using invalid characters in file names such as *? \ / : # % ∼ { }
12. The file names of records relating to recurring events should include the date and a description of the event, except where the inclusion of these elements would be incompatible with rule 3.
13. The version number of a record should be indicated in its file name by the inclusion of ‘V’ followed by the version number (e.g. V01, V03 etc.). However versioning is enabled automatically in systems such as Office 365 and One Drive for Business, making it unnecessary to duplicate this information in the file name itself. e.g. 2021-11-19_Topic_Filename-variable01

How will we keep data safe and secure?

- Delete personal & confidential details at the earliest opportunity (specify when)
- Use digital storage that require a username/password or other security feature
- Physical security (such as locked cabinet or room)
- Encrypt storage devices
- Encrypt data during transfer
- Avoid cloud services located outside EU
- Take ‘Information Security Awareness training’
- Ensure backups are also held securely

The aggregated data that is shared among project staff and partners will not contain any names, however the data being shared may still permit the identification of individuals depending on the domains being shared and may therefore constitute pseudo-anonymised data.

We also note that there is not adequate shared secure storage space at LSHTM. We will have to use our personal H drives which is suboptimal for joint working and version control.

**ARCHIVING & SHARING**

All data will be stored for 10 years.

- Files intended for sharing may be hosted in the LSHTM data repository (http://datacompass.lshtm.ac.uk) or a 3rd party repository, such as UK Data Service, ArrayExpress, Zenodo, etc.
- Internal and confidential files can be held on the LSHTM Secure Server
- Internal confidential files will be retained on Peek’s secure servers.
- LSHTM analyses will be saved on encrypted and password-protected files on LSHTM SharePoint, with access restricted to the project team. Once the project is complete these files will be moved to a secure server.
- Data presented in publications (anonymised aggregate mean attendance rates for each SES subgroup) will be published on GitHub.

Resources will be made available at the same time as findings are published in an academic journal. Once available, we will make other researchers aware that the resources exist by:

- Citing resources in future research papers, e.g. in the data access statement or reference list
- Citing resources in project reports
- Adding resources to a list of our academic outputs

The following steps will be taken to ensure that resources are easy to analyse and use in future research:

- Store resources in open file formats such as CSV, Rich Text, etc. See https://www.ukdataservice.ac.uk/manage-data/format/recommended-formats
- Designate a corresponding author / data custodian who will handle data-related questions

Conditions on access/use

| Requirement: | To be addressed by: |
| --- | --- |
| In line with the UK concordat on open research data (2016), anonymised data from this trial will be made available to bona fide research groups (evidenced via CVs and the involvement of a qualified statistician), and in line with the trial's publicly available data sharing policy, following review and approval from the trial's data monitoring committee. No reasonable request will be turned down, and the appropriate data will be made available within 1-month of receiving the request. | The PI will forward requests for data to the in-country leads in order to seek the relevant permissions. We will respond to any bona fide request within 28 days. |
| There may be multiple levels of permission required in-country before data can be shared, including national ministry of health approval and local implementation partner approval |  |

**RESOURCING**

With respect to costs of resources, we have adequate funding within the Wellcome project grant. The data is collected through active live Peek powered programmes where funding and resources is already provided for data collection and data security.

